# Maternal and child genetic liability for smoking and caffeine consumption and child mental health: An intergenerational genetic risk score analysis in the ALSPAC cohort

**DOI:** 10.1101/2020.09.07.20189837

**Authors:** Laura Schellhas, Elis Haan, Kayleigh E Easey, Robyn E Wootton, Hannah M Sallis, Gemma C Sharp, Marcus R Munafò, Luisa Zuccolo

## Abstract

**Background and aims:** Previous studies suggest an association between maternal tobacco and caffeine consumption during and outside of pregnancy and offspring mental health. We aimed to separate effects of the maternal environment (intrauterine or postnatal) from pleiotropic genetic effects.

**Design:** Secondary analysis of a longitudinal study. We 1) validated smoking and caffeine genetic risk scores (GRS) derived from published GWAS for use during pregnancy, 2) compared estimated effects of maternal and offspring GRS on childhood mental health outcomes, and 3) tested associations between maternal and offspring GRS on their respective outcomes.

**Setting:** We used data from a longitudinal birth cohort study from England, the Avon Longitudinal Study of Parents and Children (ALSPAC).

**Participants:** Our sample included 7921 mothers and 7964 offspring.

**Measurements:** Mental health and non-mental health phenotypes were derived from questionnaires and clinical assessments: 79 maternal phenotypes assessed during and outside of pregnancy, and 71 offspring phenotypes assessed in childhood (<10 years) and adolescence (11-18 years).

**Findings:** The maternal smoking and caffeine GRS were associated with maternal smoking and caffeine consumption during pregnancy (2^nd^ trimester: P_smoking_ = 3.0×10^−7^, P_caffeine_ = 3.28×10^−5^). Both the maternal and offspring smoking GRS showed evidence of association with reduced childhood anxiety symptoms (β_maternal_ = -0.033; β_offspring_= -0.031) and increased conduct disorder symptoms (β_maternal_= 0.024; β_offspring_= 0.030), after correcting for multiple testing. Finally, the maternal and offspring smoking GRS were associated with phenotypes related to sensation seeking behaviours in mothers and adolescence (e.g., increased symptoms of externalising disorders, extraversion, and monotony avoidance). The caffeine GRS showed weaker evidence for associations with mental health outcomes.

**Conclusions:** We did not find strong evidence that maternal smoking and caffeine genetic risk scores (GRS) have a causal effect on offspring mental health outcomes. Our results confirm that the smoking GRS also captures liability for sensation seeking personality traits.

## INTRODUCTION

Smoking and caffeine consumption often co-occur (1) and are associated with mental health problems and other substance use behaviours (2,3). There is some evidence that smoking is a causal risk factor for mental health problems, such as depression and schizophrenia (4,5); however, the relationship between caffeine and mental health is less clear, and possibly difficult to disentangle from smoking as the two often co-occur (3,6). In addition to associations between smoking, caffeine and mental health outcomes within individuals, observational research suggests that prenatal maternal consumption of tobacco and caffeine could have an intergenerational effect on offspring’s mental health (7–10).

Using conventional epidemiological methods alone, it is difficult to ascertain whether prenatal tobacco and caffeine exposure causally affect offspring mental health outcomes (11,12). Not only do mothers and offspring share a similar environment (such as socio-economic position), they also share, on average, 50% of their segregating genetic variation. Due to this shared genetic and environmental confounding it is difficult to disentangle the effect of maternal substance use on offspring mental health from those of offspring’s own substance use.

The association between maternal prenatal smoking and internalising problems in children is less extensively researched compared to associations with externalising problems, and existing evidence is mixed (9,13–15). Many studies report a positive association between prenatal smoking and offspring’s externalising problems (7,16–18), which could reflect a potential intrauterine effect of smoking. However, results vary when adopting different methods to account for shared environmental and genetic confounders (16). For example, studies using negative controls designs and sibling comparisons have found inconclusive evidence for a causal intrauterine effect (16,17,19– 21). In fact, study designs adjusting for shared genetic factors between mother and offspring have concluded that genetic factors explain associations between maternal prenatal smoking and externalising problems in offspring (22). This literature highlights the complexity of the nature of associations between pregnancy exposures and offspring mental health, and the importance of disentangling shared genetic and environmental confounders to understand whether a true causal effect exists.

Using genetic risk scores (GRS) as proxies for smoking or caffeine consumption can, in principle, reduce bias from confounding (23). However, when investigating intergenerational effects this approach may lead to spurious results for several reasons (24). First, the genetic variants used in the GRS have mostly been identified and validated in non-pregnant adult populations and thus might not predict behaviour during pregnancy (24–26). Second, offspring’s own smoking or caffeine consumption may confound associations because mothers pass on their genetic predisposition for smoking or caffeine consumption to their children. Consequently, when offspring’s mental health outcomes are assessed at an age where offspring are likely to have started smoking or drinking caffeine themselves, offspring’s own consumption may cause offspring’s mental health problems. Third, an association between maternal GRS and offspring mental health outcomes may reflect a shared genetic liability for smoking or caffeine consumption and mental health outcomes (pleiotropy) instead of a causal effect of the exposure. Given the shared genetics between parents and offspring, intergenerational GRS analyses should control for both offspring GRS and paternal GRS to avoid collider bias, but often it is not possible due to the limited availability of data on mothers, fathers and offspring, and limited sample size, in many cohort studies (14).

In this study, we aimed to elucidate the effects of maternal prenatal smoking and caffeine consumption on offspring mental health, using data from a multi-generational cohort study from England, the Avon Longitudinal Study of Parents and Children (ALSPAC) (27). We had two specific aims: 1) to validate the smoking and caffeine GRS during pregnancy (in mothers) and adolescence (in offspring) and 2) to estimate the effect of maternal smoking and caffeine consumption on offspring mental health. The second aim was achieved by first estimating the association between maternal smoking and caffeine GRS with offspring mental health outcomes during childhood (before age 10 years when children are unlikely to start smoking or consuming higher level of caffeine themselves; childhood GRS analysis, Figure 1), and then comparing the effect of mothers GRS and offspring GRS on offspring mental health to disentangle pleiotropic from potential causal associations (intergenerational GRS analysis, Figure 1).

**Figure 1.**
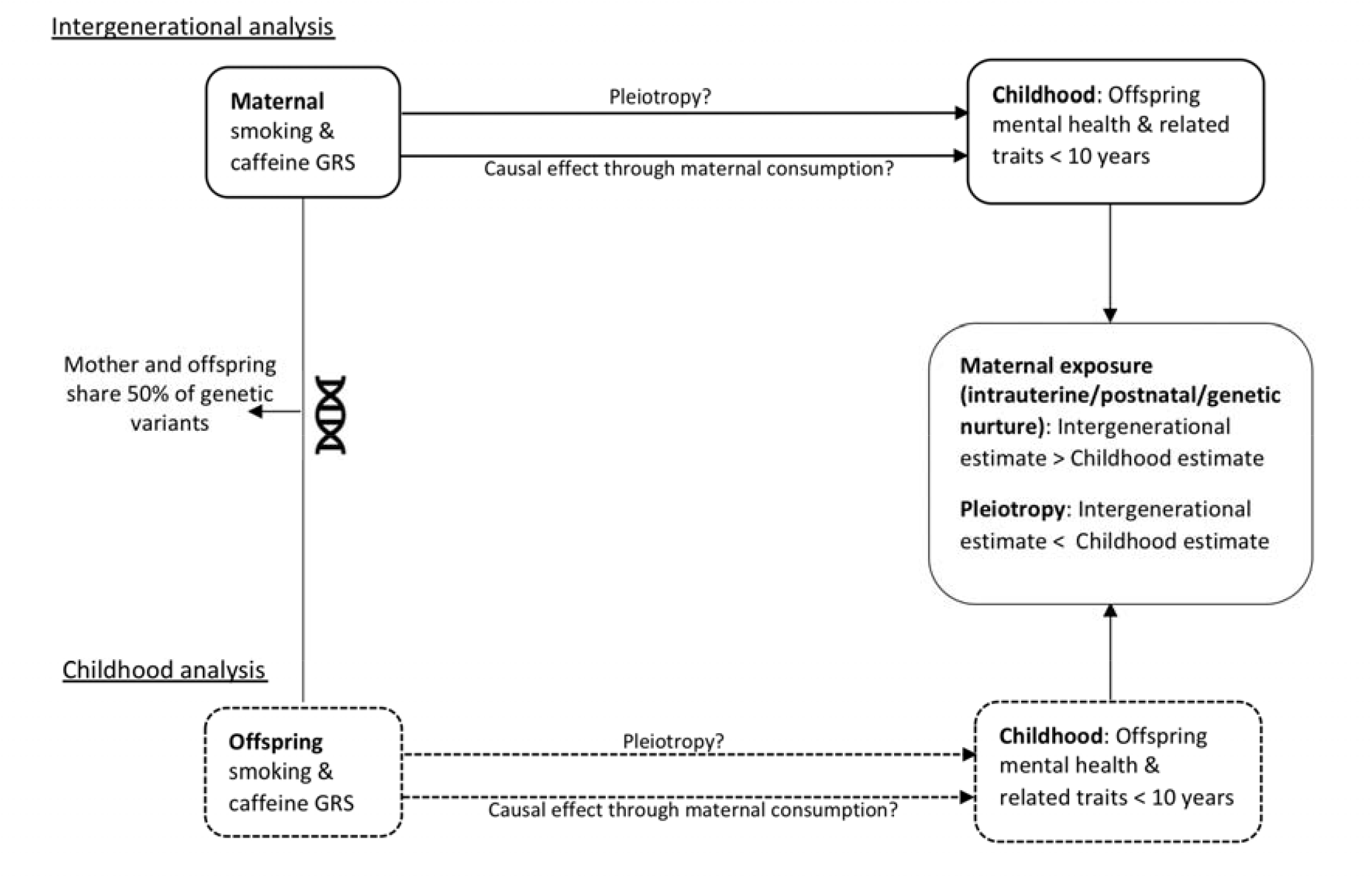
Analyses of Aim 2: Comparison of intergenerational and childhood analysis results to disentangle maternal environmental from pleiotropic effects. *Note*. Design overview of the analyses of aim 2, which compares the intergenerational analysis (top) and the childhood analysis (bottom). A larger effect estimate in the intergenerational compared to the childhood analysis would reflect a causal effect of caffeine/smoke exposure through the maternal environment (intrauterine/postnatal/genetic nurture). A larger effect estimate in the childhood compared to the intergenerational analysis would reflect pleiotropic association of the genetic risk scores with mental health.

## METHODS

### Design

A visual overview of the study design can be found in Figure 1. Given the shared genetic material between mothers and offspring, we expect pleiotropic associations to be reflected by a larger estimated effect of the offspring GRS on offspring mental health, compared to the estimated effect of the maternal GRS (childhood GRS analysis). Following the same reasoning, a larger estimated effect of the maternal GRS on offspring mental health (relative to the estimated effect of the offspring GRS) would provide more evidence to support a causal effect of maternal behaviour on offspring mental health (intergenerational GRS analysis).

### Study population

The Avon Longitudinal Study of Parents and Children (ALSPAC) is a prospective longitudinal cohort study where the initial number of pregnancies enrolled is 14,541 and of these initial pregnancies, there were a total of 14,676 fetuses, resulting in 14,062 live births and 13,988 children who were alive at 1 year of age. When the oldest children were approximately 7 years of age, an attempt was made to bolster the initial sample with eligible cases who had failed to join the study originally, resulting in an additional 913 children being enrolled. The total sample size for analyses using any data collected after the age of seven is therefore 15,454 pregnancies, resulting in 15,589 fetuses. Of these 14,901 were alive at 1 year of age (27–29). The ALSPAC study was approved by the ALSPAC Ethics and Law Committee and the Local Research Ethics Committees and informed consent for the use of data collected via questionnaires and clinics was obtained from participants. The study website contains details of all the data that is available through a fully searchable data dictionary and variable search tool: http://www.bristol.ac.uk/alspac/researchers/our-data/

### Phenotype data

Mental health phenotypes were selected from questionnaires and clinical assessments. Besides mental health phenotypes, some non-mental health phenotypes were also included, that were selected based on evidence in the literature indicating high comorbidity with mental health problems (e.g., alcohol, cannabis, other drugs, personality, body mass index, sleep, socio-economic variables). To validate the GRS, we derived phenotypes to describe caffeine consumption and smoking behaviours. Offspring assessment points were grouped into ‘childhood’ (age 7-11 years) and ‘adolescence’ (age 12-18 years). Maternal assessment points were grouped into ‘during pregnancy’ (8, 18 and 32 weeks of gestation) and ‘outside of pregnancy’, which included phenotypes assessed pre- and/or post-pregnancy. Outcomes assessed within the first four years after pregnancy were excluded, as the transition to parenthood may influence mental health temporarily (30) and mothers may be more likely to be pregnant again. In total we included 71 phenotypes for offspring (childhood and adolescence) and 79 phenotypes for mothers (during and outside of pregnancy). Table 1 gives an overview of phenotypes included in the intergenerational and childhood GRS analyses across time-points. A complete list of phenotypes is given in Supplementary Table S1. More details about the phenotype selection and assessment can be found in Supplementary Methods.

**Table 1.** Availability of phenotypes included in the intergenerational and childhood analyses across sub-populations.

| Measures | Timepoints |  |  |  |
| --- | --- | --- | --- | --- |
|  | Offspring |  | Mothers |  |
|  | Childhood (age<10) | Adolescence (age 12-18) | Outside of pregnancy (pre/post-pregnancy) | During pregnancy |
| <u>Mental health</u> |  |  |  |  |
| ***Emotional problems*** |  |  |  |  |
| Depression symptoms | x | x | x | x |
| Anxiety symptoms | x | x | x | x |
| Specific phobia | x | x |  |  |
| ***Behavioural problems*** |  |  |  |  |
| Oppositional defiant disorder symptoms | x | x |  |  |
| Conduct disorder symptoms | x | x | Personality measures (extraversion, anger, impulsivity) |  |
| ADHD symptoms | x | x |  |  |
| Total behavioural difficulties | x | x |  |  |
| ***Neuro-developmental*** |  |  |  |  |
| Autism (lifetime diagnosis) |  | x |  |  |
| <u>Other</u> |  |  |  |  |
| Handedness (negative control) | x |  |  |  |
| IQ | x | x | Only education & SES | Only education & SES |
| Number of stressful life events | x | x | x | x |
| BMI | x | x | x | Only Image perception and physical activity |
| Sleep initiation | x | x |  |  |
| Sleep maintenance | x | x |  |  |
| Hours of sleep (duration) | x | x | x |  |
| Overall caffeine intake | x | x | x | x |

### Genetic risk scores (GRS)

In ALSPAC, genome-wide SNP data were available for 8,237 children and 8,196 mothers (detailed information about genotyping can be found in Supplementary methods). After removing individuals who withdrew their consent or did not pass quality control, GRS could be generated for 7,964 children and 7921 mothers (see Supplementary methods for more details (31)). The GWAS and Sequencing Consortium of Alcohol and Nicotine use (GSCAN, N = 1.2 million (25)) identified 378 single nucleotide-polymorphisms (SNPs) associated with smoking initiation that were conditionally independent at the genome-wide significance level (P< 5×10^−8^). Smoking initiation was defined as being an ‘ever’ vs. ‘never’ smoker where an ‘ever’ smoker had to have either smoked 100 cigarettes in their lifetime and/or smoked regularly every day for at least a month. Of the 378 genome-wide significant SNPs, 356 were available in ALSPAC (25). Considering that smoking is a complex behaviour, of which initiation is only one part, we also generated a GRS for lifetime smoking. The lifetime smoking score also captures smoking heaviness (as well as smoking duration and cessation) but is derived in the entire population comprising both smokers and non-smokers and therefore is more suitable for use in unstratified samples (4). The GWAS of lifetime smoking based on the UK Biobank data (N=462,690) identified 126 independent loci (p<5×10^−8^), which were all available in ALSPAC. The Coffee and Caffeine Genetics Consortium found 8 SNPs to be independently associated with cups of coffee consumed per day at the genome-wide level of significance (N = 91,462 (26)), which were all available in ALSPAC. These SNPs have also been found to be associated with caffeine use from other caffeinated beverages (32,33).

We created weighted genetic risk scores using independent genome wide significant hits (P<5×10^−8^) and their effect estimates as reported in the discovery GWAS for each of our exposures. These GRS were derived using Plink v1.9 and standardised prior to use in analyses. As our GRS were based on discovery GWAS that only report independent variants (4,25,26), clumping or pruning were not necessary (34).

### Statistical analysis

The statistical analysis plan for this secondary analysis of study data was not pre-registered and the results should be considered exploratory.

All analyses were performed using Stata v15 (35). The following linear and logistic regression analyses were conducted to test associations with the smoking and caffeine GRS: 1) maternal GRS with smoking and caffeine phenotypes in mothers during pregnancy to validate the GRS (Aim 1); 2) maternal and offspring GRS with childhood phenotypes (<10 years) for investigating intergenerational effects (Figure 1) (Aim 2); and supplementary analyses 3) maternal and offspring GRS with their own phenotypes in mothers (during and outside of pregnancy) and offspring (adolescence) to confirm GRS associations with relevant substance use behaviours as a positive control and gain more information about mental health associations at later times in development. Analyses were adjusted for age, offspring sex, and the first 10 ancestry-informative principal components based on the ALSPAC genome-wide data. We restricted our sample to singletons or one individual from a twin pair and to individuals of European ancestry. The maximum sample size available in childhood was 6,156 (4,974 in adolescence) and 7,269 during pregnancy (7,199 outside of pregnancy). To avoid limiting our sample size further, and to reduce the risk of selection bias, we did not restrict our analyses to only mother-offspring pairs with complete genotype data. We compared mother-offspring pairs where either mother or offspring have genotype data but not both with respect to smoking, caffeine and socio-demographic variables. This comparison is shown in Supplementary Table S2.

### Multiple testing

Multiple testing was accounted for by running Monte Carlo permutation testing with 1000 repetitions. These p-values are presented in the text. We also compared these results with a more stringent Bonferroni correction. However, given the high degree of correlation between our phenotypes, this correction is likely to be overly conservative. Evidence for association was considered strongest for phenotypes that also survived Bonferroni correction (all results are available in the Supplementary material).

## RESULTS

### Maternal smoking and caffeine consumption

In our sample, 51% of mothers reported having ever smoked a cigarette in their lifetime and 25% reported smoking during the first trimester of pregnancy. Mothers’ median caffeine consumption outside of pregnancy (97 months post-pregnancy) was 168 milligrams of caffeine a day (mg/day; interquartile range (IQR): 108 to 252). During pregnancy, mothers reported lower caffeine consumption with a median of 138 mg/day (IQR: 81 to 215) during the 2^nd^ trimester and 135 mg/day (IQR: 71 to 216) during the 3^rd^ trimester. Compared to mothers who did not report smoking, mother who smoked reported consistently more caffeine consumption during (2^nd^ trimester: 64 mg/day more caffeine, 3^rd^ trimester: 75 mg/day more caffeine) and outside of pregnancy (8 years post-pregnancy: 30 mg/day more caffeine).

### Validation of GRS during pregnancy

The GRS for smoking initiation and lifetime smoking were positively associated with maternal smoking phenotypes during pregnancy and explained 1-5% of variance in smoking phenotypes during and outside of pregnancy (Table 2 and Supplementary Table S3). The GRS for caffeine consumption was positively associated with total caffeine and caffeinated tea and coffee consumption during pregnancy, but not with cola consumption (Table 3). The caffeine GRS explained 0.2-0.4% of variance in caffeine phenotypes during pregnancy and 0.2-1% of variance outside of pregnancy (Table 2).

**Table 2.** Associations between smoking initiation genetic risk scores (GRS) and smoking phenotypes in mothers (during and outside of pregnancy) and offspring in adolescence.

|  | Phenotype | Effect estimate | Effect size* | 95% CI | p-value | Sample size | Adj. R<sup>2**</sup> |
| --- | --- | --- | --- | --- | --- | --- | --- |
| **Mothers** |  |  |  |  |  |  |  |
| **outside of pregnancy** | Mother has ever smoked | OR | 1.40 | 1.33, 1.48 | 1.24x10<sup>-8</sup> | 7194 | 0.03 |
|  | Number of cigarettes smoked before pregnancy | Beta | 0.15 | 0.08, 0.22 | 3.81x10<sup>-5</sup> | 3426 | 0.05 |
| **Pregnancy – 18 weeks gestation** | Tobacco smoked in 1<sup>st</sup> three months of pregnancy | OR | 1.35 | 1.23, 1.44 | 3.0x10<sup>-7</sup> | 7237 | 0.05 |
|  | Mother cut down smoking | OR | 1.33 | 1.25, 1.42 | 5.89x10<sup>-7</sup> | 7269 | 0.03 |
|  | Mother stopped smoking during pregnancy | OR | 0.98 | 0.88, 1.11 | 0.771 | 1863 | 0.01 |
| **Offspring** |  |  |  |  |  |  |  |
| **Adolescence– 14 years** | Smoked at age 14 years | OR | 1.18 | 1.09, 1.28 | 6.50x10<sup>-4</sup> | 4145 | 0.03 |
|  | Smoked more than 20 cigarettes | OR | 1.19 | 1.03, 1.38 | 0.024 | 1058 | 0.03 |
|  | Age 1<sup>st</sup> smoked a cigarette | Beta | 0.001 | -0.04, 0.04 | 0.953 | 1064 | 0.01 |
| **Adolescence– 18 years** | Ever smoked a whole cigarette | OR | 1.26 | 1.15, 1.37 | 1.09x10<sup>-4</sup> | 2402 | 0.02 |
|  | Number of cigarettes smoked in lifetime | Beta | 0.19 | 0.10, 0.2 | 4.24x10<sup>-5</sup> | 1144 | 0.01 |

\* Reflects the average change in the outcome that is associated with a one standard deviation increase in the GRS. For binary outcomes, this will be the odds ratio (e.g., Mother's odds of ever smoking are 1.4 times compared to not smoking), for continuous outcomes it represents the average unit change (e.g., 0.15 cigarettes smoked). \*\* For the logistic regression models pseudo R<sup>2</sup> is reported.

**Table 3.**
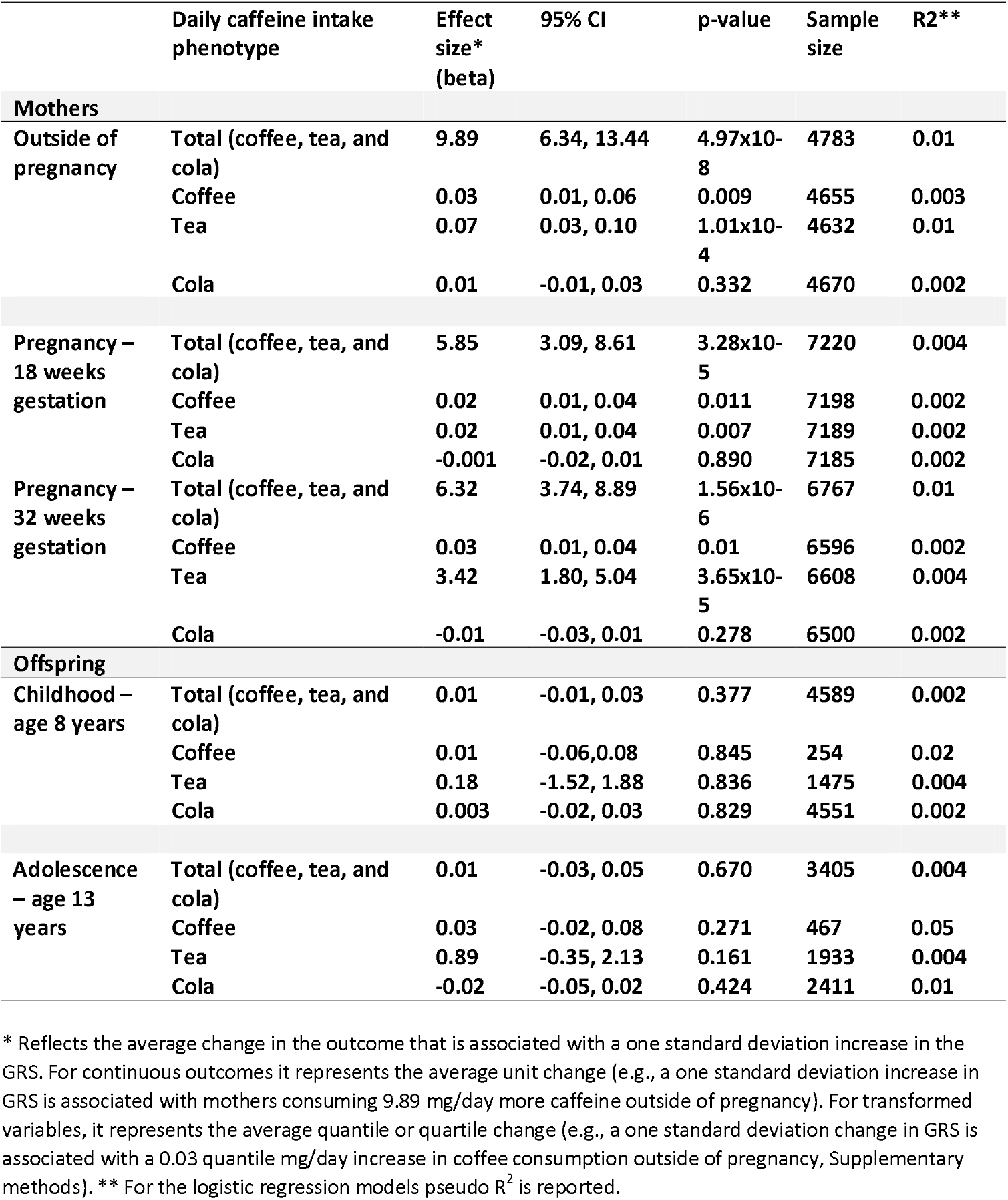
Associations between caffeine genetic risk scores (GRS) and daily caffeine intake in mothers (during and outside of pregnancy), and offspring (age 8 and 13 years).

### Comparison of intergenerational and childhood smoking initiation GRS analyses

#### Intergenerational GRS analyses

Of 17 childhood mental health phenotypes, the strongest evidence of association was observed <u>for</u> reduced anxiety symptoms (P_perm_=0.002) and increased conduct disorder symptoms (P_perm_=0.021). Of the non-mental health phenotypes, the strongest associations were found for lower IQ (P_perm_=0.02), higher overall caffeine consumption (P_perm_= <0.001) and BMI (P_perm_=0.001) as well as the likelihood of being left-handed (P_perm_=0.012), which was included as a negative control phenotype (because we would not expect a causal intrauterine effect of maternal smoking or caffeine on handedness). The only associations to survive Bonferroni correction (P <0.003) were that of maternal smoking GRS with offspring’s anxiety symptoms and offspring’s caffeine consumption (Figure 2, Table 4).

**Figure 2.**
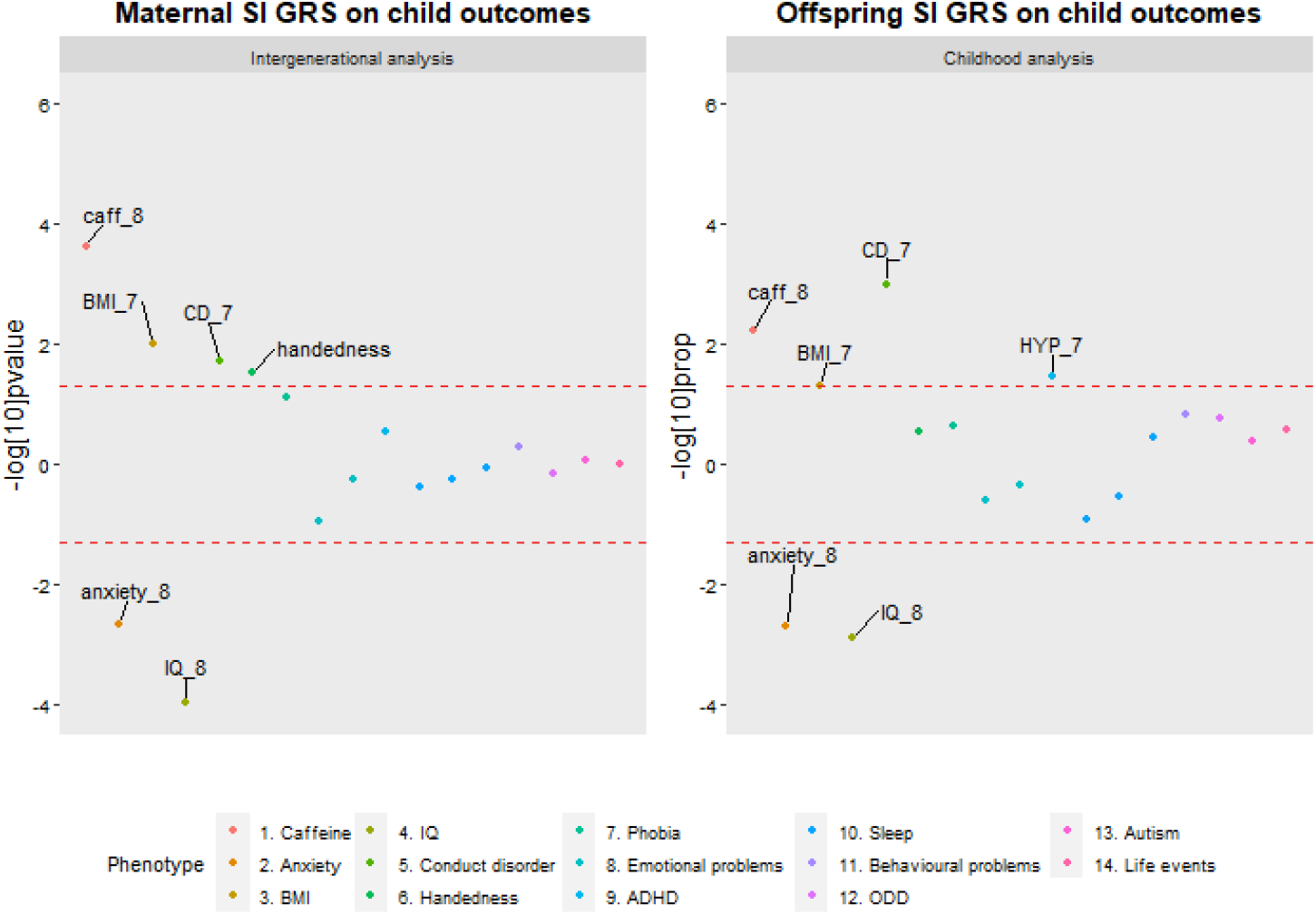
Comparison of phenotype associations with the smoking initiation genetic risk scores (SI GRS) in the intergenerational and childhood analysis. *Note*. Points outside the lines had a permutation corrected p-value < 0.05. Points above the upper line represent positive associations and points below the lower line represent negative associations. caff_8 = Total caffeine consumption at age 8. BMI_7 = BMI at age 7. CD = Conduct Disorder at age 7. anxiety_8 = Anxiety at age 8. IQ_8 = IQ at age 8. HYP_7 = Hyperactivity at age 8.

**Table 4.** Associations between the maternal and offspring smoking initiation GRS and offspring phenotypes <10 years.

|  |  | Intergenerational analyses |  |  |  |  |  | Childhood analyses |  |  |  |  |  |
| --- | --- | --- | --- | --- | --- | --- | --- | --- | --- | --- | --- | --- | --- |
|  |  | Regression analyses |  |  | Permutation testing |  |  | Regression analyses |  |  | Permutation testing |  |  |
| Phenotype | Effect estimate | Effect size | 95% CI | P-value | 95% CI | P-value | Sample size | Effect size | 95% CI | P-value | 95% CI | P-value | Sample size |
| 1. Total caffeine | Beta | 0.045 | 0.021, 0.068 | <0.001 | <0.001, 0.004 | <0.001 | 4067 | 0.032 | 0.010, 0.055 | 0.005 | 0.002, 0.013 | 0.006 | 4589 |
| 2. Anxiety | Beta | -0.033 | -0.053, -0.012 | 0.002 | <0.001, 0.007 | 0.002 | 4993 | -0.031 | -0.051, -0.010 | 0.003 | <0.001, 0.007 | 0.002 | 5355 |
| 3. BMI | Beta | 0.076 | 0.018, 0.135 | 0.010 | 0.007, 0.022 | 0.013 | 5032 | 0.050 | <0.001, 0.101 | 0.051 | 0.036, 0.063 | 0.048 | 5799 |
| 4. IQ | Beta | -0.592 | -1.049, -0.134 | 0.011 | 0.009, 0.026 | 0.016 | 4675 | -0.735 | -1.183, -0.287 | 0.001 | <0.001, 0.004 | <0.001 | 5295 |
| 5. Conduct disorder | Beta | 0.024 | 0.004, 0.044 | 0.019 | 0.013, 0.032 | 0.021 | 5012 | 0.030 | 0.012, 0.049 | 0.001 | <0.001, 0.006 | 0.001 | 5326 |
| 6. Handedness | OR | 1.114 | 1.012, 1.225 | 0.030 | 0.006, 0.021 | 0.012 | 4849 | 1.045 | 0.954, 1.145 | 0.315 | 0.263, 0.320 | 0.291 | 5403 |
| 7. Specific phobia | OR | 1.322 | 0.964, 1.813 | 0.078 | 0.042, 0.071 | 0.055 | 5100 | 1.182 | 0.881, 1.587 | 0.241 | 0.199, 0.252 | 0.225 | 5470 |
| 8. Emotional problems | Beta | -0.016 | -0.037, 0.004 | 0.117 | 0.106, 0.148 | 0.126 | 5139 | -0.011 | -0.031, 0.009 | 0.267 | 0.236, 0.291 | 0.263 | 5459 |
| 9. ADHD | Beta | 0.016 | -0.013, 0.045 | 0.277 | 0.232, 0.287 | 0.259 | 4916 | 0.030 | 0.003, 0.058 | 0.030 | 0.024, 0.047 | 0.034 | 5219 |
| 10. Sleep duration | Beta | -0.009 | -0.033, 0.014 | 0.426 | 0.392, 0.454 | 0.423 | 5127 | -0.019 | -0.042, 0.004 | 0.106 | 0.107, 0.149 | 0.127 | 5443 |
| 11. Behavioural difficulties | Beta | 0.010 | -0.021, 0.041 | 0.522 | 0.482, 0.544 | 0.513 | 5133 | 0.022 | -0.008, 0.051 | 0.152 | 0.130, 0.176 | 0.152 | 5452 |
| 12. Depression | Beta | -0.006 | -0.027, 0.015 | 0.557 | 0.524, 0.586 | 0.555 | 4885 | -0.007 | -0.027, 0.012 | 0.466 | 0.442, 0.504 | 0.473 | 5434 |
| 13. Sleep maintenance | OR | 0.983 | 0.919, 1.051 | 0.589 | 0.534, 0.596 | 0.565 | 5127 | 0.973 | 0.913, 1.038 | 0.383 | 0.313, 0.372 | 0.342 | 5448 |
| 14. ODD | Beta | -0.004 | -0.024, 0.016 | 0.700 | 0.683, 0.740 | 0.712 | 4943 | 0.015 | -0.005, 0.034 | 0.148 | 0.146, 0.194 | 0.169 | 5319 |
| 15. Autism | OR | 1.027 | 0.722, 1.460 | 0.874 | 0.860, 0.901 | 0.882 | 5975 | 1.153 | 0.803, 1.654 | 0.411 | 0.380, 0.442 | 0.411 | 6156 |
| 16. Sleep initiation | OR | 0.995 | 0.934, 1.061 | 0.874 | 0.827, 0.873 | 0.851 | 5150 | 0.971 | 0.913, 1.032 | 0.309 | 0.269, 0.326 | 0.297 | 5476 |
| 17. Life events | Beta | <0.001 | -0.018, 0.019 | 0.996 | 0.991, 0.999 | 0.997 | 5167 | 0.010 | -0.008, 0.028 | 0.271 | 0.237, 0.292 | 0.264 | 5493 |

*Note.* The intergenerational analysis represents offspring phenotypes <10 years regressed on maternal GRS. The childhood analysis represents offspring phenotypes <10 years regressed on offspring GRS.

#### Childhood GRS analyses

As observed in the intergenerational analysis, there was some evidence for an association with reduced anxiety problems (P_perm_=0.002) and increased conduct disorder symptoms (P_perm_=0.001). In contrast to the intergenerational analysis, there was some evidence for an association with ADHD symptoms (P_perm_=0.034). The strongest non-mental health associations of the intergenerational analysis were replicated using the offspring smoking GRS (lower IQ, P_perm_< 0.001; increased caffeine consumption: P_perm_=0.048) with the exception of left-handedness (P_perm_=0.291; Figure 3). Only <u>the</u> associations with IQ and conduct disorder symptoms survived Bonferroni correction of P <0.003 (Figure2, Table 4).

**Figure 3.**
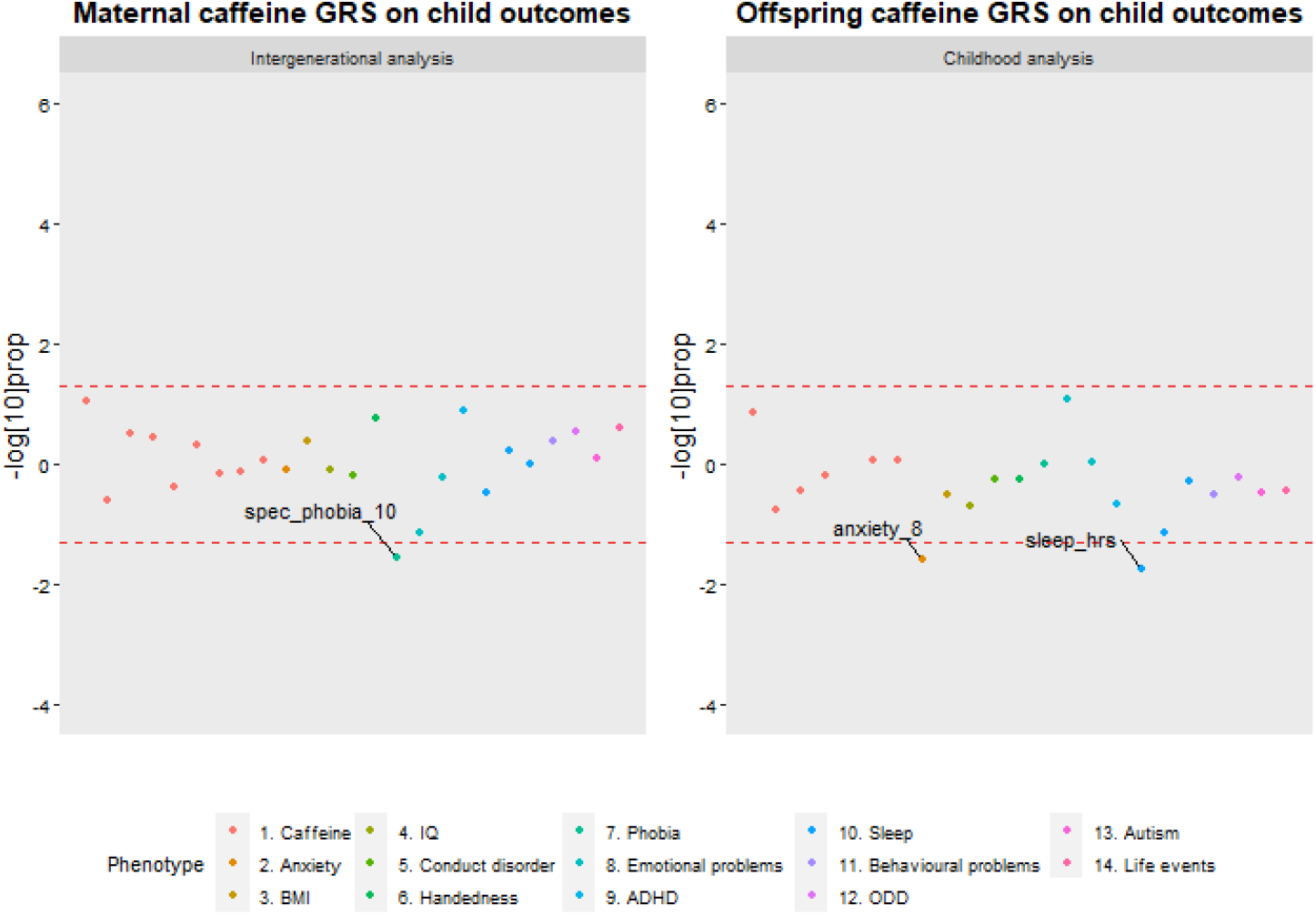
Comparison of phenotype associations with the caffeine genetic risk scores (GRS) in the intergenerational and childhood analysis. *Note*. Points outside the lines had a permutation corrected p-value < 0.05. Points above the upper line represent positive associations and points below the lower line represent negative associations. spec_phobia_10 = specific phobias at age 10. anxiety_8 = Anxiety at age 8. sleep_hrs = Sleep_hrs = Sleep duration in hours at age 7.

The results using lifetime smoking GRS were largely consistent. Only the associations with offspring’s IQ, ODD and total behavioural difficulties survived Bonferroni correction (Supplementary Table S4).

### Comparison of intergenerational and childhood caffeine GRS analyses

#### Intergenerational GRS analyses

Given that offspring’s caffeine GRS was not robustly associated with caffeine consumption in childhood (Table 3), we were able to use the results of the childhood analysis as a way of assessing pleiotropy, despite some children already consuming low levels of caffeine at this age. Of the 17 childhood phenotypes, the strongest mental health association was observed with decreased risk for specific phobias in offspring (P_perm_=0.028; Figure 3). There was no evidence for associations with any of the non-mental health phenotypes.

#### Childhood GRS analyses

In contrast to the intergenerational analysis, there was no evidence for an association with specific phobias (P_perm_=0.998) but some evidence for an association with reduced general anxiety symptoms (P_perm_=0.026). <u>The</u> strongest association amongst the non-mental health phenotypes was observed with fewer hours of sleep in term-time (P_perm_=0.018), (Figure 3 and Table 5). None of the associations of the intergenerational and childhood analyses for caffeine survived Bonferroni correction.

**Table 5.** Associations between maternal and offspring caffeine GRS and offspring phenotypes <10 years.

|  |  | Intergenerational analyses |  |  |  |  |  | Childhood analyses |  |  |  |  |  |
| --- | --- | --- | --- | --- | --- | --- | --- | --- | --- | --- | --- | --- | --- |
|  |  | Regression analyses |  |  | Permutation testing |  |  | Regression analyses |  |  | Permutation testing |  |  |
| Phenotype | Effect estimate | Effect size | 95% CI | P-value | 95% CI | P-value | Sample size | Effect size | 95% CI | P-value | 95% CI | P-value | Sample size |
| 1. Specific phobia | OR | 0.724 | 0.519, 1.012 | 0.057 | 0.019, 0.040 | 0.028 | 5100 | 0.999 | 0.723, 1.381 | 0.997 | 0.993, 1.000 | 0.998 | 4900 |
| 2. Depression | Beta | -0.017 | -0.039, 0.002 | 0.075 | 0.056, 0.089 | 0.071 | 4885 | -0.017 | -0.037, 0.002 | 0.081 | 0.055, 0.088 | 0.070 | 5434 |
| 3. ADHD | Beta | -0.018 | -0.008, 0.050 | 0.161 | 0.110, 0.152 | 0.130 | 4916 | -0.018 | -0.046, 0.010 | 0.206 | 0.199, 0.251 | 0.224 | 5219 |
| 4. Handedness | OR | 1.064 | 0.968, 1.169 | 0.178 | 0.143, 0.189 | 0.165 | 4849 | 0.980 | 0.897, 1.070 | 0.624 | 0.548, 0.610 | 0.579 | 5399 |
| 5. Life events | Beta | -0.008 | -0.007, 0.030 | 0.228 | 0.217, 0.271 | 0.243 | 5167 | -0.008 | -0.027, 0.010 | 0.366 | 0.329, 0.390 | 0.359 | 5493 |
| 6. ODD | Beta | 0.002 | -0.032, 0.008 | 0.240 | 0.257, 0.314 | 0.285 | 4943 | 0.002 | -0.018, 0.022 | 0.829 | 0.583, 0.644 | 0.614 | 5319 |
| 7. Sleep initiation | OR | 0.972 | 0.913, 1.036 | 0.352 | 0.316, 0.375 | 0.345 | 5150 | 0.951 | 0.895, 1.010 | 0.094 | 0.059, 0.092 | 0.074 | 5476 |
| 8. Total caffeine | Beta | 0.008 | -0.015, 0.032 | 0.490 | 0.444, 0.506 | 0.475 | 4067 | 0.010 | -0.012, 0.032 | 0.377 | 0.349, 0.410 | 0.379 | 4589 |
| 9. BMI | Beta | 0.025 | -0.033, 0.084 | 0.387 | 0.364, 0.425 | 0.394 | 5032 | 0.025 | -0.027, 0.077 | 0.348 | 0.297, 0.356 | 0.326 | 5799 |
| 10. Behavioural difficulties | Beta | -0.015 | -0.019, 0.043 | 0.441 | 0.369, 0.431 | 0.400 | 5133 | -0.015 | -0.044, 0.015 | 0.324 | 0.294, 0.353 | 0.323 | 5452 |
| 11. Emotional problems | Beta | 0.001 | -0.014, 0.027 | 0.538 | 0.569, 0.631 | 0.600 | 5139 | 0.001 | -0.018, 0.021 | 0.883 | 0.881, 0.919 | 0.901 | 5459 |
| 12. Sleep duration | Beta | -0.026 | -0.030, 0.017 | 0.577 | 0.544, 0.606 | 0.575 | 5127 | -0.026 | -0.048, -0.004 | 0.018 | 0.011, 0.028 | 0.018 | 5443 |
| 13. CD | Beta | -0.006 | -0.024, 0.015 | 0.624 | 0.627, 0.686 | 0.657 | 5012 | -0.006 | -0.024, 0.013 | 0.563 | 0.541, 0.603 | 0.572 | 5326 |
| 14. Autism | OR | 1.052 | 0.758, 1.461 | 0.742 | 0.733, 0.787 | 0.761 | 5975 | 0.850 | 0.603, 1.199 | 0.326 | 0.307, 0.366 | 0.336 | 6156 |
| 15. IQ | Beta | 0.276 | -0.521, 0.390 | 0.778 | 0.766, 0.817 | 0.792 | 4675 | 0.276 | -0.155, 0.707 | 0.209 | 0.183, 0.234 | 0.208 | 5290 |
| 16. Anxiety | Beta | -0.022 | -0.023, 0.019 | 0.849 | 0.805, 0.853 | 0.830 | 4993 | -0.022 | -0.042, -0.002 | 0.029 | 0.017, 0.038 | 0.026 | 5355 |
| 17. Sleep maintenance | OR | 1.001 | 0.936, 1.071 | 0.970 | 0.947, 0.972 | 0.961 | 5127 | 0.983 | 0.922, 1.048 | 0.573 | 0.517, 0.579 | 0.548 | 5488 |

*Note.* The intergenerational analysis represents offspring phenotypes <10 years regressed on maternal GRS. The childhood analysis represents offspring phenotypes <10 years regressed on offspring GRS.

### Smoking and caffeine GRS analyses with phenotypes during and outside of pregnancy and during adolescence

The GRS for smoking were associated with these behaviours outside of pregnancy and during adolescence (Table 2). The caffeine GRS was associated with caffeine consumption outside of pregnancy (except for cola consumption) but not during adolescence (Table 3). The strongest evidence for associations with the smoking GRS was found for binge drinking, increased caffeine consumption, BMI and extraverted personality traits in mothers (Bonferroni threshold = 0.002) and more externalising problems and extraversion, increased BMI, and lower IQ in adolescence (Bonferroni threshold = 0.001). A detailed description of these results can be found in the Supplementary Material and Supplementary Tables S5-S7. None of the caffeine GRS associations survived Bonferroni correction (Supplementary Table S6).

## DISCUSSION

In this study we aimed to disentangle possible causal associations of maternal smoking and caffeine consumption, with a particular focus on the prenatal period, on offspring mental health from pleiotropic associations. Our results showed that the maternal smoking and caffeine GRS are valid predictors of smoking and caffeine consumption from tea and coffee during pregnancy. The maternal and offspring smoking initiation GRS were associated with various mental health traits and other substance use behaviours across different time points in life. In particular, we observed associations of the maternal and offspring smoking initiation GRS with sensation-seeking traits across development, such as less anxiety and increased externalising problems in childhood, an extroverted personality type, more externalising problems and alcohol consumption in adolescence, as well as higher expression of anger, more monotony avoidance outside of pregnancy and alcohol consumption during and outside of pregnancy. We found few associations between the maternal and offspring caffeine GRS and offspring mental health outcomes. Critically, our results indicate that the associations found between the maternal smoking and caffeine GRS and offspring mental health outcomes are likely due to pleiotropic effects, rather than acting through the maternal intrauterine environment.

The literature supports our findings of pleiotropic associations between the maternal and offspring smoking GRS and sensation-seeking personality traits. Previous studies found that adolescence who smoke have more externalising problems, higher impulsivity and novelty-seeking type of behaviours (36) and that children with lower cognitive abilities have more behavioural problems and are more likely to initiate smoking themselves (37,38). There is evidence for shared genetic factors influencing smoking behaviours, externalising problems and novelty seeking type of behaviours (39,40), as well as educational attainment (41). However, some studies argue that the effect from the maternal postnatal environment (such as parenting behaviours) and maternal mental health cannot be dismissed even after accounting for genetic effects (42,43). We found some evidence that the maternal smoking GRS is associated with maternal depression during and outside of pregnancy, which could (partly) explain the association we observed between the maternal smoking GRS and offspring externalising problems. A study adopting a similar design to the present one, examining associations between maternal and offspring GRS for increased alcohol consumption and maternal and offspring mental health (44), also found an association between maternal alcohol use and maternal depression during pregnancy but no evidence for an association with maternal alcohol GRS and externalising problems in offspring. Even though this requires further testing, it could provide some initial evidence that the association between the maternal smoking GRS and offspring externalising problems is more likely to be pleiotropic than confounded by maternal depression. Further, other studies suggest that the genetic instrument for smoking initiation may not only measure smoking behaviour but also capture novelty-seeking and impulsive behaviours even when only using genome-wide significant SNPs (41,45–47). In addition, GSCAN summary statistics for smoking initiation showed a strong genetic correlation with ADHD and risk tolerance behaviour, which could make pleiotropic effects more likely (25). Taken together with the existing literature, our findings support the notion that these observed associations with maternal smoking initiation GRS are likely explained by shared genetic liability in mothers and offspring.

We did not find strong evidence for intergenerational effects between the maternal caffeine GRS and offspring mental health outcomes in childhood. The associations we observed between maternal caffeine GRS and decreased likelihood of binge drinking, reduced caffeine consumption and lower socioeconomic status during pregnancy, as well as the offspring caffeine GRS and higher GCSE exam grades during adolescence stand-in contrast to a study in the UK Biobank where the caffeine GRS was positively associated with alcohol consumption outside of pregnancy and not associated with social class (32). Therefore, these findings should be interpreted with caution, as they might be unique to the ALSPAC sample in terms of participants’ sociodemographic characteristics or false positives. Although these results could be due to yet unexplained forms of bias, it is also possible that the caffeine GRS is capturing underlying personality/socio-behavioural profiles with far reaching consequences for health and wellbeing, which deserves further investigation.

### Strengths and Limitations

A major strength of this study was the exploration of exposure-outcome associations at time points in life other than adulthood. Further, the validation of genetic variants discovered in non-pregnant female and male populations, as proxies during pregnancy, is vital for future investigation of intrauterine effects of the exposures (24). Lastly, the intergenerational comparison of associations of the maternal smoking and caffeine GRS with childhood mental health outcomes, that are likely to be free of confounding through offspring’s own substance consumption enabled us to disentangle potential pleiotropic and environmental effects on mental health.

The following limitations should also be considered. First, the limited sample size (in the context of genetic association studies) likely resulted in low statistical power to detect small effects. Second, we were restricted to phenotypes as assessed in ALSPAC, and the comparison of related phenotypes was not similar across development (e.g., ADHD/conduct disorder in childhood with extraversion & anger personality traits in mothers outside of pregnancy). Third, many mental health phenotypes in childhood were based on maternal report, which may not accurately reflect offspring’s mental health problems (48,49) but rather mothers own mental health status (50,51). Fourth, we constructed GRS for smoking initiation based on the latest GWAS that included ALSPAC mothers (25). Due to the sample overlap, the true strength of explored associations might be smaller than we reported. However, given the small contribution of data from ALSPAC (∼1%) to a total sample size of 1.2 million, the risk of bias is likely negligible. Fifth, to make the smoking GRS specific to our exposure of interest we based our GRS on genome-wide significant SNPs only, yet the smoking GRS still showed associations with some alcohol phenotypes. We checked the correlations between the alcohol, smoking and caffeine GRS, which were low (Supplementary Table S8). However, because of the phenotypic associations with alcohol consumption, we cannot rule out that associations observed with the maternal smoking GRS are cofounded by maternal alcohol consumption. Still, this is unlikely to affect our results because we did not find evidence for potential causal effects, and previous research by Easey and colleagues observed no associations in intergenerational analyses between maternal alcohol GRS and offspring mental health outcomes (44). Sixth, as our dataset included phenotypes from later time points and we relied on participants whose genotype data was available, it is possible that our findings are subject to selection bias (31,52). Last, the comparison of the intergenerational and childhood GRS analyses was based on transmitted alleles and therefore an indirect effect of maternal non-transmitted alleles on offspring sensation-seeking traits through genetic nurturing cannot be ruled out (53).

### Future research

Future studies investigating the effects on mental health using the smoking initiation GRS might consider accounting for sensation seeking personality traits. Further, future research should aim to differentiate effects of smoke exposure through the intrauterine and postnatal environment, explore non-linear effects of the smoking and caffeine GRS, and investigate a potential interaction of smoking and caffeine consumption during pregnancy on offspring mental health (54). More analyses exploiting paternal data would be helpful to understand the effect of smoking and caffeine consumption on offspring mental health outcomes. For instance, studies with paternal genotype data could help to differentiate whether observed effects are due to intrauterine or postnatal exposure, through conducting negative control comparisons of prenatal associations of maternal and paternal substance use.

## Conclusion

In conclusion, our study validated the application of the smoking initiation, lifetime smoking and caffeine GRS for research investigating intrauterine exposures to smoking and caffeinated coffee and tea. Further, we found stronger evidence for pleiotropic rather than causal effects of maternal smoking and caffeine consumption on offspring mental health. Given the current study’s limitations, particularly its limited statistical power, these findings should be replicated in independent samples using more refined methods for pleiotropy detection and corrections.

## Data Availability

Data is available through application to the research executive of ALPSAC. Please note that the study website contains details of all the data that is available through a fully searchable data dictionary and variable search tool.

http://www.bristol.ac.uk/alspac/researchers/our-data/

## Declarations of interest

None.

## Acknowledgements

We are extremely grateful to all the families who took part in this study, the midwives for their help in recruiting them, and the whole ALSPAC team, which includes interviewers, computer and laboratory technicians, clerical workers, research scientists, volunteers, managers, receptionists and nurses. The UK Medical Research Council and Wellcome (Grant ref: 217065/Z/19/Z) and the University of Bristol provide core support for ALSPAC. This publication is the work of the authors and Laura Schellhas and Elis Haan will serve as guarantors for the contents of this paper.

## Funding

This research was performed in the UK Medical Research Council Integrative Epidemiology Unit (grant number: MC_UU_00011/7) and also supported by the National Institute for Health Research (NIHR) Bristol Biomedical Research Centre at University Hospitals Bristol NHS Foundation Trust and the University of Bristol. The MRC also funded KEE’s PhD studentship. LZ was supported by a UK Medical Research Council fellowship (grant number G0902144). GCS was supported by the Medical Research Council [New Investigator Research Grant, MR/S009310/1] and the European Joint Programming Initiative “A Healthy Diet for a Healthy Life” (JPI HDHL, NutriPROGRAM project, UK MRC MR/S036520/1].

This research was also conducted as part of the CAPICE (Childhood and Adolescence Psychopathology: unravelling the complex etiology by a large Interdisciplinary Collaboration in Europe) project, funded by the European Union’s Horizon 2020 research and innovation programme, Marie Sklodowska Curie Actions – MSCA-ITN-2016 – Innovative Training Networks under grant agreement number 721567. This study was supported by the NIHR Biomedical Research Centre at the University Hospitals Bristol NHS Foundation Trust and the University of Bristol. The views expressed in this publication are those of the authors and not necessarily those of the NHS, the National Institute for Health Research or the Department of Health and Social Care.

The views expressed in this publication are those of the authors and not necessarily those of the National Health Service, the National Institute for Health Research or the Department of Health. A comprehensive list of grant funding is available on the ALSPAC website (http://www.bristol.ac.uk/alspac/external/documents/grant-acknowledgements.pdf). GWAS data was generated by Sample Logistics and Genotyping Facilities at Wellcome Sanger Institute and LabCorp (Laboratory Corporation of America) using support from 23andMe.

## Notes

### Competing Interest Statement

The authors have declared no competing interest.

### Author Declarations

Ethics approval for the study was obtained from the ALSPAC Ethics and Law Committee and the Local Research Ethics Committees.

### Summary of Updates

This version is an update of introduction and genetic risk scores section, includes a design section and provides more clarity on results section. Supplemental file updated.

