## supplementary material for "Maternal and child genetic liability for smoking and caffeine consumption and child mental health: An intergenerational genetic risk score analysis in the ALSPAC cohort"

**SUPPLEMENTARY INFORMATION**

**Supplementary Methods**

**Phenotype selection and coding**

Phenotype assessment

Phenotypes that correlated highly, only the one with the larger sample size was included. If some phenotypes were assessed at close time points (e.g. at age 14 and age 16) but only showed a low to moderate association, both time points were included. Continuous phenotypes that substantially deviated from a normal distribution were transformed into quantiles and If validated cut-off score was available, binary phenotypes were derived. Zero inflation was accounted for by transforming continuous phenotypes with more than 20% of zero values into 3 quantiles (0, <median, >median). An observation was classified as an outlier if it fell outside 1.5 times the interquartile range^[1]^.

Caffeine phenotypes

Caffeine phenotypes (coffee, tea, cola) were transformed based on their caffeine content. In ALSPAC, caffeine content for a cup of tea was 27mg, for a cup of coffee 57mg and for a can of cola (330ml) 20mg of caffeine^[2]^. Total caffeine consumption was computed by summing up each drink. Extreme outliers, such as consuming more than 28 cups of coffee and tea per day were removed.

**Genotype data**

In ALSPAC, genome-wide data was available for 8237 children and 8196 mothers. The ALSPAC children genotype data has been generated using the Illumina HumanHap550 quad chip genotyping platforms by 23andme subcontracting the Wellcome Trust Sanger Institute, Cambridge, UK and the Laboratory Corporation of America, Burlington, NC, US. Mothers were genotyped using Illumina human660w quad array at the Centre National Genotypage (CNG) and genotypes were called with Illumina GenomeStudio. Quality control filtering was done with the PLINK (v1.07) software. SNPs with a minor allele frequency of < 1%, call rate < 95% and Hardy-Weinberg equilibrium (HWE) *P* < 5E-7 were removed. Both offspring and maternal genotype data have been jointly imputed to the 1000 genomes reference panel (version 1, Phase 3, Dec 2013 Release).

**Supplementary Analyses**

**Associations between the maternal and offspring GRS and own mental health during and outside of pregnancy (mothers) and adolescence (offspring).**

We assessed associations between the maternal and offspring GRS and own mental health during and outside of pregnancy (mothers) and adolescence (offspring) to explore whether the associations were pleiotropic or suggestive of possible causal relationships. Further, comparison of the magnitude of effects across childhood, adolescence and adulthood could give indication about the persistence of pleiotropic effects across development and the effect on mental health through maternal and offspring’s own behaviour.

Maternal smoking initiation GRS and outcomes during and outside of pregnancy.

Amongst the mental health phenotypes, we found consistent evidence for the maternal smoking GRS being associated with increased depressive symptoms during and outside of pregnancy.

After applying Bonferroni correction (P<0.002), the strongest evidence was found for associations with lower education, higher caffeine consumption and binge drinking during and outside of pregnancy, as well as with higher BMI, lower image perception and personality traits such as anger and monotony avoidance outside of pregnancy (Supplementary Table S4).

Offspring smoking initiation GRS and own outcomes during adolescence

Consistent with the results using the childhood mental health phenotypes, there was some evidence for positive associations with externalising problems in adolescence (conduct disorder, oppositional defiant disorder, ADHD). However, there was very little evidence of association with anxiety symptoms during adolescence.

After applying Bonferroni correction (P<0.001), the strongest evidence was found for associations with increased conduct disorder symptoms, higher BMI, lower IQ, more extraverted personality traits and alcohol consumption (Supplementary Table S4).

The results using the lifetime smoking GRS were largely consistent. The strongest evidence was found for associations with education, social class, caffeine consumption, BMI, and anger phenotypes in mothers during and outside of pregnancy and conduct disorder, extraversion, and IQ phenotypes in adolescence (Supplementary Table S5).

Maternal caffeine GRS and own outcomes during and outside of pregnancy

There was some evidence for an association with decreased likelihood of having had schizophrenia diagnosis outside of pregnancy but no other mental health outcomes during or outside of pregnancy. Amongst the non-mental health phenotypes some evidence was observed for associations with less substance use during pregnancy (higher likelihood of reducing caffeine consumption, decreased likelihood to binge drink), as well as evidence for an association with lower socio-economic position. None of these associations survived Bonferroni correction (Supplementary Table S6).

Offspring caffeine GRS and own mental health outcomes during adolescence

There was only some evidence for the offspring caffeine GRS being associated with higher GCSE exam grades during adolescence but none of the mental health or substance use phenotypes. None of the associations survived Bonferroni correction (Supplementary Table S8).

**Supplementary Table S1. List of phenotypes included in the study**

| **Phenotype** | **Assessment instrument** | **Timepoint** |
| --- | --- | --- |
| **Offspring: Children** |  |  |
| **Mental health** |  |  |
| ADHD symptoms (categorical) | SDQ^[3]^ | 6.7 years |
| Conduct disorders symptoms (categorical) | SDQ | 6.7 years |
| Oppositional-defiant disorder symptoms (categorical) | DAWBA^[4]^ | 7.5 years |
| SMFQ score (categorical) | SMFQ^[5]^ | 9.5 years |
| SDQ emotional symptoms score (categorical) | SDQ | 6.8 years |
| General anxiety symptoms score (categorical) | DAWBA | 7.5 years |
| Total behavioural difficulties score (categorical) | SDQ | 6.7 years |
| Specific phobia clinical diagnosis (binary) | DAWBA | 9.5 years |
| Autism diagnosis (binary) | Derived combining multiple measures^[6]^ | 9 years |
| **Non-mental health** |  |  |
| Sleep duration in hours | Maternal report | 6.7 years |
| Number of life events (categorical) | Life events inventory (maternal report) | 6.7 years |
| Problems with sleep initiation in past year (binary) | Maternal report | 6.7 years |
| Problems with sleep maintenance in past year (binary) | Maternal report | 6.7 years |
| IQ total score | WISC^[7]^ | 8 years |
| Body mass Index |  | 7 years |
| Right or left-handed (binary) | Child completed | 11 years |
| *Caffeine* |  |  |
| Total mg/day caffeine from tea, cola, coffee | Maternal report | 8 years |
| Child drinks caffeinated tea (binary) | Maternal report | 8 years |
| Child drinks caffeinated coffee (binary) | Maternal report | 8 years |
| **Offspring: Adolescents** |  |  |
| **Mental health** |  |  |
| ADHD symptoms (categorical) | SDQ | 16.6 years |
| Conduct disorder symptoms (categorical) | SDQ | 16.6 years |
| Oppositional-defiant disorder symptoms (categorical) | DAWBA | 15.5 years |
| Sleep duration | Child completed | 15.5 years |
| Number of life events (categorical) | Life events inventory (child self-report) | 16.5 years |
| Depressive symptoms – Sum of all the 5 depression symptoms (categorical) | CIS-R^[8]^ | 18 years |
| Depression symptoms total score (categorical) | MFQ^[9]^ | 17.5 years |
| Depression symptoms score (categorical) | MFQ | 14 years |
| PTSD (self-report 6-band computer prediction, binary) | DAWBA | 15 years |
| Self-harming behaviour with suicidal intent (binary) | Derived from multiple measures^[10]^ | 15 years |
| Emotional symptoms score (categorical) | SDQ | 16.5 years |
| Anxiety score (categorical) | CIS-R | 17.1 years |
| Phobias: Phobia symptom score (categorical) | CIS-R | 17.1 years |
| Total behavioural difficulties score (categorical) | SDQ | 16.5 years |
| Ever treated for an eating disorder (binary) | Child completed | 13 years |
| Psychosis positive symptoms (categorical) | Psychosis interview | 12 years |
| Psychosis negative symptoms score | PLIKS^[11]^ | 16.5 years |
| Psychosis positive symptoms (categorical) | Psychosis interview | 18 years |
| Ever treated for an eating disorder (binary) | Child completed | 16 years |
| **Non-mental health** |  |  |
| Big-5 personality traits: Extraversion | IPIP^[12]^ | 13 years |
| Big-5 personality traits: Agreeableness | IPIP | 13 years |
| Big-5 personality traits: Conscientiousness | IPIP | 13 years |
| Big-5 personality traits: Emotional Stability (neuroticism) | IPIP | 13 years |
| Big-5 personality traits: Intellect | IPIP | 13 years |
| Maintaining sleep: Number of times young person usually wakes up at night (categorical) | Child completed | 15 years |
| Initiating sleep: Average time (minutes) YP takes to fall asleep per week (categorical) | Child completed | 15 years |
| Frequency respondent did any exercise during the past year (categorical) | Child completed | 14 years |
| GCSE grades A-C (binary) | Child completed | 18 years |
| GCSE grades D-G (binary) | Child completed | 18 years |
| BMI |  | 17 years |
| IQ total score | WASI^[13]^ | 15.5 years |
| **Substance use** |  |  |
| *Alcohol* |  |  |
| AUDIT: Frequency young person has a drink containing alcohol (Continuous) | Child Self-report | 17.1 years |
| Level of risk identified by alcohol use disorders identification test | AUDIT^[14]^ | 17.1 years |
| Frequency had 6+ drinks on one occasion | Child Self-report | 17.1 years |
| No. full drinks needed to feel tipsy/have buzz over last 3 months (categorical) | Child Self-report | 17.1 years |
| Number of alcoholic drinks on a typical day (categorical) | Child Self-report | 18 years |
| Total score of AUDIT test (categorical) | AUDIT | 18 years |
| Number of times had whole drink in the past 6 months (categorical) | Self-report | 12 years |
| Number of drinks took to feel different after first 5 times drinking (categorical) | Self-report | 12 years |
| Number of times had 3+ drinks in one day (categorical) | Self-report | 12 years |
| *Tobacco* |  |  |
| Age of respondent when first smoked a cigarette | Self-report | 14 years |
| Age when respondent smoked first whole cigarette (years) | Self-report | 18 years |
| Number of cigarettes respondent smoked altogether in lifetime (categorical) | Self-report | 18 years |
| Frequency young person smokes cannabis (categorical) | Self-report | 16.5 years |
| Respondent has smoked a cigarette (including roll-ups) (binary) | Self-report | 14 years |
| Total number of cigarettes that the respondent has smoked (binary) | Self-report | 14 years |
| Respondent has ever smoked a whole cigarette (including roll-ups) (binary) | Self-report | 18 years |
| Young person has ever tried cannabis (binary) | Self-report | 16.5 years |
| *Caffeine* |  |  |
| Total mg/day caffeine from tea, coffee, cola (categorical) | Maternal report | 13 years |
| Tea mg/day caffeine teenager | Maternal report | 13 years |
| Coffee mg/day caffeine teenager (categorical) | Maternal report | 13 years |
| Cola mg/day caffeine teenager (categorical) | Maternal report | 13 years |
| **Mothers during pregnancy** |  |  |
| **Mental health** |  |  |
| Depression symptoms (binary) | EPDS^[15]^ | 18 weeks gest |
| Depression symptoms (binary) | EPDS | 32 weeks gest |
| Hypersensitivity to interpersonal rejection | IPMS^[16]^ | 18 weeks gest |
| Anxiety symptoms (binary) | CCEI^[17]^ |  |
| **Non-mental health** |  |  |
| Number of life events mother experienced in pregnancy (categorical) | Life events Inventory | 18 weeks gest |
| Image perception score during pregnancy | Self-report | 18 weeks gest |
| Image perception change from before to during pregnancy | Self-report | 18 weeks gest |
| Your reactions to becoming a parent (categorical) | Self-report | 18 weeks gest |
| Activity level compared with other pregnant women (categorical) | Self-report | 32 weeks gest |
| Physical activity (binary) | Self-report | 32 weeks gest |
| Vomited in first three months of pregnancy (binary) | Self-report | 18 weeks gest |
| Social class based on occupation (categorical) | Self-report | 32 weeks gest |
| Mothers highest education in pregnancy (categorical) | Self-report | 32 weeks gest |
| **Substance use** |  |  |
| *Alcohol* |  |  |
| Alcohol: binging (categorical) | Self-report | 18 weeks gest |
| Alcohol per week | Self-report | 32 weeks gest |
| Alcohol: binging (categorical) | Self-report | 32 weeks gest |
| *Tobacco* |  |  |
| Smoking first three months in pregnancy (binary) | Self-report | 18 weeks gest |
| Ever smoked during pregnancy (binary) | Self-report | 8 weeks gest |
| Stopped smoking during pregnancy (binary) | Self-report | 8 weeks gest |
| Cut down smoking during pregnancy (binary) | Self-report | 8 weeks gest |
| *Caffeine* |  |  |
| Total mg/day caffeine pregnancy | Self-report | 18 weeks gest |
| Tea mg/day caffeine pregnancy (categorical) | Self-report | 18 weeks gest |
| Coffee mg/day caffeine pregnancy (categorical) | Self-report | 18 weeks gest |
| Cola mg/day caffeine pregnancy (categorical) | Self-report | 18 weeks gest |
| Total mg/day caffeine pregnancy | Self-report | 32 weeks gest |
| Tea mg/day caffeine pregnancy | Self-report | 32 weeks gest |
| Coffee mg/day caffeine pregnancy (categorical) | Self-report | 32 weeks gest |
| Cola mg/day caffeine pregnancy (categorical) | Self-report | 32 weeks gest |
| Consumed more caffeine during pregnancy (binary) | Self-report | 8 weeks gest |
| Never has been drinking caffeine (binary) | Self-report | 8 weeks gest |
| Did not change caffeine consumption during pregnancy (binary) | Self-report | 8 weeks gest |
| Reduced caffeine consumption during pregnancy (binary) | Self-report | 8 weeks gest |
| Never drank tea vs. drinking tea (binary) | Self-report | 8 weeks gest |
| Stopped drinking tea during pregnancy (binary) | Self-report | 8 weeks gest |
| Reduced tea consumption during pregnancy (binary) | Self-report | 8 weeks gest |
| Craved or had more tea during pregnancy (binary) | Self-report | 8 weeks gest |
| Never drank coffee vs. drinking coffee (binary) | Self-report | 8 weeks gest |
| Stopped drinking coffee during pregnancy (binary) | Self-report | 8 weeks gest |
| Reduced coffee consumption during pregnancy | Self-report | 8 weeks gest |
| Craved or had more coffee during pregnancy (binary) | Self-report | 8 weeks gest |
| Never drank cola vs. drinking cola (binary) | Self-report | 8 weeks gest |
| Stopped drinking cola during pregnancy (binary) | Self-report | 8 weeks gest |
| Reduced cola consumption during pregnancy (binary) | Self-report | 8 weeks gest |
| Craved or had more cola during pregnancy (binary) | Self-report | 8 weeks gest |
| Cutting down cola consumption during pregnancy (binary) | Self-report | 8 weeks gest |
| *Other substances* |  |  |
| Hard drugs (binary) | Self-report | 18 weeks gest |
| Cannabis first three months in pregnancy (binary) | Self-report | 8 weeks gest |
| **Mothers outside of pregnancy** | | |
| **Mental health** |  |  |
| Anxiety symptoms (binary) | CCEI | 11 years (child age) |
| Depression symptoms (binary) | EPDS | 11 years |
| Ever had bulimia (binary) | Self-report | 12 weeks gest |
| Ever had drug addiction (binary) | Self-report | 12 weeks gest |
| Ever had alcoholism (binary) | Self-report | 12 weeks gest |
| Ever had schizophrenia (binary) | Self-report | 12 weeks gest |
| Ever had anorexia nervosa (binary) | Self-report | 12 weeks gest |
| Ever had severe depression (binary) | Self-report | 12 weeks gest |
| Ever had other psychiatric problem (binary) | Self-report | 12 weeks gest |
| Image perception 3 months before pregnancy (categorical) | Self-report | 18 weeks gest |
| **Non-mental health** |  |  |
| Number of life events mums (categorical) | Life-events inventory (self-report) | 11 years |
| Impulsivity trait | KSP^[18]^ | 9 years |
| Monotony avoidance trait | KSP | 9 years |
| Anger trait | KSP | 9 years |
| Suspicion trait | KSP | 9 years |
| Detachment trait | KSP | 9 years |
| Social class based on occupation (categorical) | Self-report | 4 years |
| Mothers highest educational qualifications (categorical) | Self-report | 5 years |
| BMI mothers | Self-report | 12 weeks gest |
| Mother participates in physical activity (binary) | Self-report | 18 years |
| **Substance use** |  |  |
| *Tobacco* |  |  |
| Mother has ever been smoker (binary) | Self-report | 18 weeks gest |
| Number of cigarettes mother smoked before pregnancy | Self-report | 18 weeks gest |
| Number of cigarettes mother has smoked last 2 weeks | Self-report | 8 years |
| *Caffeine* |  |  |
| Daily caffeine intake from cola (mg) (categorical) | Self-report | 8 years |
| Daily caffeine intake from tea (mg) (categorical) | Self-report | 8 years |
| Daily caffeine intake from coffee (mg) (categorical) | Self-report | 8 years |
| Mothers daily caffeine intake through tea, coffee & cola (including persons with missing 1 or 2 drinks) | Self-report | 8 years |
| *Alcohol* |  |  |
| Mothers total alcohol units daily (categorical) | Self-report | 8 years |
| Mothers pre-pregnancy drinking (never/ever) (binary) | Self-report | 18 weeks gest |
| Number of days in past month that mother had at least 4 units of alcohol (categorical) | Self-report | 5 years |
| Mothers total alcohol units daily (categorical) | Self-report | 4 years |
| AUDIT score in mothers (based on risk level) (categorical) | AUDIT | 18 years |

**Supplementary Table S2. Comparison of participants with complete and partially missing genotype data**

*Two-sample t-tests (two-sided):*

|  | **Participants with genetic data*** | **N** | **Mean (SE)** | **P diff** |
| --- | --- | --- | --- | --- |
| **Caffeine consumption during pregnancy (mg/day)** | Mothers and offspring | 4918 | 160 (1.76) | 0.0002 |
|  | Mothers or offspring | 4778 | 151 (1.60) |  |
| **Social class*** | Mothers and offspring | 4,052 | 1.98 (0.02) | <0.001 |
|  | Mothers or offspring | 3,733 | 1.74 (0.02) |  |
| **Maternal education**** | Mothers and offspring | 3,935 | 2.1 (0.02) | <0.001 |
|  | Mothers or offspring | 3,353 | 2.4 (0.02) |  |
| **Maternal age (years)** | Mothers and offspring | 4,788 | 27.57 (0.07) | <0.001 |
|  | Mothers or offspring | 4,938 | 28.63 (0.07) |  |

*Note. Mothers and offspring refers to mother-offspring pairs that both have genotype data in ALSPAC. Mothers or offspring refers to mother-offspring pairs where either mother or offspring have genotype data but not both. *Social class levels are based on individual’s occupation where classes I to V stands for occupations: I – professional; II – managerial and technical; III – skilled non-manual and manual; IV – partly-skilled; V – unskilled*

*** 4 categories: 0 = none or CSE , 1 = vocational, 2 = O-level, 3 = A-level, 4 = degree (CSE reflects to the certificate of secondary education which is available for both academic and vocational subjects. O level is equivalent to grades D and E and A level is equivalent to grades A to C after GCSE (General Certificate of Secondary Education) examination. Degree level reflects to higher education diploma*

*Chi-Square test:* *Χ*^2^(1) = 69.57, P < 0.001

|  | **Participants with genetic data*** | |  |
| --- | --- | --- | --- |
| **Maternal smoking during the 1^st^ trimester of pregnancy (yes/no)** | **Mothers and offspring** | **Mothers or offspring** | **Total N** |
| **Yes** | N = 924 | N = 1304 | 2,228 |
| **No** | N = 3864 | N = 3634 | 7,498 |
| **Total N** | 4,788 | 4,938 | 9,726 |

*Note. *Mothers and offspring refers to mother-offspring pairs that both have genotype data in ALSPAC. Mothers or offspring refers to mother-offspring pairs where either mother or offspring have genotype data but not both.*

**Supplementary Table S3.** **Associations between the lifetime smoking GRS and smoking phenotypes in mothers during and outside of pregnancy and adolescents**

| **Phenotype** | **Effect estimate** | **Effect size*** | **95% CI** | **p-value** | **Sample size** | **Adj. R^2^**** |
| --- | --- | --- | --- | --- | --- | --- |
| **Mothers during pregnancy** | | | | | |  |
| Tobacco smoked in 1^st^ three months of pregnancy | OR | 1.235 | 1.159, 1.315 | 9.41x10^-6^ | 7237 | 0.04 |
| Mother cut down tobacco consumption | OR | 1.168 | 1.097, 1.244 | <0.001 | 7269 | 0.02 |
| Mother stopped smoking during pregnancy | OR | 0.871 | 0.775, 0.979 | 0.024 | 1863 | 0.01 |
| **Mothers outside of pregnancy** | | | | | |  |
| Mother has ever smoked | OR | 1.147 | 1.089, 1.209 | <0.001 | 7194 | 0.01 |
| Number of cigarettes mother smoked before pregnancy | Beta | 0.194 | 0.124, 0.264 | 5.27x10^-8^ | 3426 | 0.05 |
| **Offspring: Adolescents** | | | | | |  |
| Smoked age 14 years | OR | 1.117 | 1.033, 1.208 | 0.009 | 4145 | 0.03 |
| Smoked more than 20 cigarettes age 14 | OR | 1.156 | 0.995, 1.342 | 0.057 | 1058 | 0.01 |
| Age 1^st^ smoked a cigarette (asked age 14) | Beta | -0.052 | -0.096, -0.009 | 0.019 | 1064 | 0.01 |
| Ever smoked a whole cigarette age 18 | OR | 1.130 | 1.035, 1.233 | 0.010 | 2402 | 0.01 |
| Number of cigarettes smoked in lifetime age 18 | Beta | 0.084 | -0.006, 0.174 | 0.069 | 1144 | 0.002 |

* Reflects the average change in the outcome that is associated with a one standard deviation increase in the GRS. For binary outcomes, this will be the odds ratio (e.g. Mother’s odds of ever smoking are 1.147 times compared to not smoking), for continuous outcomes it represents the average unit change (e.g. 0.775 cigarettes smoked). ** For the logistic regression models pseudo R^2^ is reported.

**Supplementary table S4. Associations between maternal and offspring lifetime smoking GRS and offspring phenotypes <10 years**

|  | | **Intergenerational analyses** | | | | | | **Childhood analyses** | | | | | |
| --- | --- | --- | --- | --- | --- | --- | --- | --- | --- | --- | --- | --- | --- |
|  |  | **Regression analyses** | | | **Permutation testing** | | | **Regression analyses** | | | **Permutation testing** | | |
| **Phenotype** | **Effect estimate** | **Effect size** | **95% CI** | **P-value** | **95% CI** | **P-value** | **Sample size** | **Effect size** | **95% CI** | **P-value** | **95% CI** | **P-value** | **Sample size** |
| IQ | Beta | -0.742 | -1.202, -0.282 | 0.002 | <0.001, 0.007 | 0.002 | 4675 | -0.929 | -1.371, -0.488 | 3.73x10^-5^ | <0.001, 0.004 | <0.001 | 5290 |
| Conduct disorder | Beta | 0.026 | 0.007, 0.045 | 0.009 | 0.003, 0.014 | 0.007 | 5012 | 0.029 | 0.010, 0.048 | 0.003 | 0.001, 0.009 | 0.003 | 5326 |
| BMI | Beta | 0.063 | 0.007, 0.119 | 0.029 | 0.020, 0.043 | 0.030 | 5032 | 0.026 | -0.025, 0.076 | 0.316 | 0.282, 0.341 | 0.311 | 5799 |
| Total caffeine | Beta | 0.021 | -0.003, 0.045 | 0.079 | 0.063, 0.097 | 0.079 | 4067 | 0.015 | -0.007, 0.038 | 0.187 | 0.170, 0.220 | 0.194 | 4589 |
| Sleep initiation | OR | 0.950 | 0.892, 1.012 | 0.104 | 0.064, 0.099 | 0.080 | 5150 | 0.968 | 0.911, 1.029 | 0.273 | 0.203, 0.256 | 0.229 | 5476 |
| Behavioural difficulties | Beta | 0.025 | -0.005, 0.056 | 0.107 | 0.089, 0.129 | 0.108 | 5133 | 0.045 | 0.016, 0.075 | 0.003 | <0.001, 0.007 | 0.002 | 5452 |
| ADHD | Beta | 0.023 | -0.006, 0.052 | 0.117 | 0.098, 0.139 | 0.117 | 4916 | 0.037 | 0.009, 0.065 | 0.009 | 0.006, 0.020 | 0.011 | 5219 |
| Specific phobia | OR | 1.222 | 0.916, 1.631 | 0.156 | 0.179, 0.230 | 0.204 | 5100 | 0.824 | 0.628, 1.083 | 0.150 | 0.169, 0.219 | 0.193 | 5470 |
| Anxiety | Beta | -0.012 | -0.033, 0.009 | 0.256 | 0.229, 0.284 | 0.256 | 4993 | -0.014 | -0.034, 0.007 | 0.189 | 0.150, 0.198 | 0.173 | 5355 |
| Sleep duration | Beta | -0.013 | -0.036, 0.010 | 0.259 | 0.243, 0.299 | 0.270 | 5127 | 0.002 | -0.021, 0.024 | 0.878 | 0.851, 0.893 | 0.873 | 5443 |
| Sleep maintenance | OR | 1.019 | 0.952, 1.090 | 0.559 | 0.503, 0.565 | 0.534 | 5127 | 0.984 | 0.924, 1.048 | 0.594 | 0.556, 0.618 | 0.587 | 5448 |
| Autism | OR | 1.104 | 0.768, 1.589 | 0.563 | 0.512, 0.574 | 0.543 | 5975 | 1.259 | 0.838, 1.891 | 0.243 | 0.163, 0.213 | 0.187 | 6156 |
| ODD | Beta | 0.006 | -0.014, 0.026 | 0.574 | 0.557, 0.619 | 0.588 | 4943 | 0.031 | 0.012, 0.051 | 0.002 | <0.001, 0.004 | <0.001 | 5319 |
| Emotional problems | Beta | -0.005 | -0.025, 0.016 | 0.656 | 0.630, 0.689 | 0.660 | 5139 | -0.012 | -0.031, 0.008 | 0.248 | 0.210, 0.264 | 0.236 | 5459 |
| Depression | Beta | -0.003 | -0.023, 0.018 | 0.809 | 0.783, 0.833 | 0.809 | 4885 | 0.010 | -0.010, 0.030 | 0.323 | 0.300, 0.359 | 0.329 | 5434 |
| Handedness | OR | 1.009 | 0.914, 1.114 | 0.846 | 0.790, 0.839 | 0.815 | 4849 | 1.006 | 0.924, 1.096 | 0.876 | 0.866, 0.906 | 0.887 | 5399 |
| Life events | Beta | -0.002 | -0.020, 0.017 | 0.853 | 0.838, 0.882 | 0.861 | 5167 | 0.014 | -0.004, 0.032 | 0.117 | 0.101, 0.143 | 0.121 | 5493 |
| Total number of outcomes tested = 17 | | | | | | | | | | | | | |

*Note.* Intergenerational analysis refers to maternal GRS predicting offspring phenotypes <10 years. Childhood analysis refers to offspring GRS predicting offspring phenotypes <10 years.

**Supplementary table S5. Associations between the maternal and offspring smoking initiation GRS and phenotypes in mothers during and outside of pregnancy and adolescence**

|  |  | **Regression analyses** | | | **Permutation testing** | | |
| --- | --- | --- | --- | --- | --- | --- | --- |
| **Phenotype** | **Effect estimate** | **Effect size** | **95% CI** | **P-value** | **95% CI** | **P-value** | **Sample size** |
| **Mothers outside of pregnancy** | | | | | | | |
| **Mental health** | | | | | | | |
| Depression symptoms | OR | 1.070 | 0.969, 1.182 | 0.161 | 0.117, 0.161 | 0.138 | 4725 |
| Anxiety symptoms | OR | 1.028 | 0.934, 1.131 | 0.542 | 0.515, 0.577 | 0.546 | 4740 |
| Bulimia | OR | 1.081 | 0.926, 1.261 | 0.295 | 0.319, 0.379 | 0.349 | 6799 |
| Drug addiction | OR | 0.938 | 0.594, 1.480 | 0.764 | 0.748, 0.801 | 0.775 | 6799 |
| Alcoholism | OR | 1.243 | 0.903, 1.711 | 0.163 | 0.118, 0.162 | 0.139 | 6799 |
| Schizophrenia | OR | 0.839 | 0.386, 1.825 | 0.632 | 0.585, 0.646 | 0.616 | 6799 |
| Anorexia nervosa | OR | 1.062 | 0.886, 1.272 | 0.484 | 0.429, 0.491 | 0.460 | 6799 |
| Severe depression | OR | 1.178 | 1.064, 1.303 | 0.004 | <0.001, 0.004 | <0.001 | 6799 |
| Other psychological problem | OR | 1.146 | 0.941, 1.396 | 0.157 | 0.103, 0.145 | 0.123 | 6799 |
| **Substance use** | | | | | | | |
| *Alcohol* | | | | | | | |
| Alcohol drinking before pregnancy | OR | 1.129 | 1.017, 1.253 | 0.026 | 0.006, 0.020 | 0.011 | 7199 |
| Binge drinking | Beta | 0.050 | 0.020, 0.080 | 0.001 | <0.001, 0.004 | <0.001 | 4866 |
| Daily alcohol units at child age 4 | Beta | 0.023 | 0.003, 0.044 | 0.027 | 0.018, 0.039 | 0.027 | 5680 |
| Daily alcohol units at child age 8 | Beta | -0.003 | -0.033, 0.027 | 0.838 | 0.799, 0.847 | 0.824 | 2707 |
| AUDIT score | Beta | 0.023 | 0.002, 0.045 | 0.036 | 0.034, 0.061 | 0.046 | 2424 |
| *Caffeine* | | | | | | | |
| Total caffeine consumption | Beta | 8.568 | 4.948, 12.187 | <0.001 | <0.001, 0.004 | <0.001 | 4783 |
| **Non-mental health** | | | | | | | |
| Life events | Beta | 0.021 | -0.012, 0.055 | 0.212 | 0.222, 0.277 | 0.249 | 4219 |
| Sleep duration | Beta | -0.023 | -0.049, 0.004 | 0.099 | 0.088, 0.127 | 0.106 | 1867 |
| Impulsivity personality trait | Beta | 0.072 | -0.034, 0.177 | 0.183 | 0.147, 0.195 | 0.170 | 4847 |
| Monotony avoidance personality trait | Beta | 0.242 | 0.099, 0.386 | 0.001 | <0.001, 0.006 | 0.001 | 4794 |
| Anger personality trait | Beta | 0.341 | 0.207, 0.475 | <0.001 | <0.001, 0.004 | <0.001 | 4769 |
| Suspicion personality trait | Beta | 0.125 | 0.016, 0.234 | 0.024 | 0.012, 0.031 | 0.020 | 4856 |
| Detachment personality trait | Beta | -0.058 | -0.169, 0.053 | 0.304 | 0.293, 0.352 | 0.322 | 4753 |
| Physical activity | OR | 0.933 | 0.858, 1.014 | 0.094 | 0.040, 0.069 | 0.053 | 2787 |
| Social class | Beta | 0.020 | -0.024, 0.064 | 0.379 | 0.422, 0.484 | 0.453 | 2906 |
| Education | Beta | -0.092 | -0.124, -0.060 | 2.19 x 10^-8^ | <0.001, 0.004 | <0.001 | 4919 |
| BMI before pregnancy | Beta | 0.210 | 0.117, 0.302 | 9.67 x 10^-6^ | <0.001, 0.004 | <0.001 | 6398 |
| Image perception before pregnancy | Beta | 0.055 | 0.029, 0.082 | 3.26x10^-5^ | <0.001, 0.004 | <0.001 | 6623 |
| Total number of outcomes tested = 27 | | | | | | | |
| **Mothers during pregnancy** | | | | | | | |
| **Mental health** | | | | | | | |
| Depression (18 wks) | OR | 1.115 | 1.028, 1.211 | 0.013 | 0.001, 0.010 | 0.004 | 6734 |
| Depression (32 wks) | OR | 1.124 | 1.039, 1.216 | 0.007 | <0.001, 0.006 | 0.001 | 6751 |
| Anxiety | OR | 0.999 | 0.915, 1.091 | 0.991 | 0.979, 0.994 | 0.988 | 6645 |
| Hypersensitivity to interpersonal rejection | Beta | -0.474 | -0.846, -0.102 | 0.012 | 0.003, 0.016 | 0.008 | 7167 |
| Feelings becoming a parent | Beta | -0.003 | -0.025, 0.018 | 0.752 | 0.724, 0.778 | 0.752 | 7165 |
| **Substance use** | | | | | | | |
| *Caffeine* | | | | | | | |
| Total caffeine (18wks) | Beta | 7.352 | 4.748, 9.957 | 3.25x10^-8^ | <0.001, 0.004 | <0.001 | 7220 |
| Total caffeine (32wks) | Beta | 6.282 | 3.693, 8.872 | 2.02x10^-6^ | <0.001, 0.004 | <0.001 | 6767 |
| *Alcohol* | | | | | | | |
| Binge drinking (18wks) | Beta | 0.043 | 0.024, 0.061 | 8.07x10^-6^ | <0.001, 0.004 | <0.001 | 7171 |
| Binge drinking (32wks) | Beta | 0.034 | 0.014, 0.054 | 0.001 | <0.001, 0.004 | <0.001 | 5324 |
| Weekly alcohol units (32wks) | Beta | 0.160 | 0.033, 0.286 | 0.013 | 0.003, 0.014 | 0.007 | 4294 |
| *Other substances* | | | | | | | |
| Cannabis use in pregnancy | OR | 1.165 | 0.977, 1.389 | 0.082 | 0.046, 0.077 | 0.060 | 6918 |
| Hard drug use in pregnancy | OR | 0.990 | 0.568, 1.726 | 0.971 | 0.947, 0.972 | 0.961 | 7147 |
| **Non-mental health** | | | | | | | |
| Education | Beta | -0.100 | -0.128, -0.071 | 1.01x10^-11^ | <0.001, 0.004 | <0.001 | 6954 |
| Social class | Beta | 0.050 | 0.023, 0.078 | 3.19x10^-4^ | <0.001, 0.004 | <0.001 | 5854 |
| Life events in pregnancy | Beta | 0.046 | 0.018, 0.074 | 0.001 | <0.001, 0.004 | <0.001 | 6744 |
| Image perception in pregnancy | Beta | 0.145 | 0.045, 0.245 | 0.005 | 0.004, 0.017 | 0.009 | 6699 |
| Image perception change | Beta | 0.077 | -0.011, 0.166 | 0.087 | 0.079, 0.117 | 0.097 | 6549 |
| Activity level compared with other pregnant women | Beta | 0.011 | -0.008, 0.029 | 0.262 | 0.226, 0.281 | 0.253 | 6611 |
| Physical activity | OR | 1.007 | 0.951, 1.066 | 0.795 | 0.764, 0.816 | 0.795 | 6767 |
| Vomiting in first three months of pregnancy | OR | 0.979 | 0.927, 1.034 | 0.418 | 0.367, 0.428 | 0.397 | 6797 |
| Sleep problems 18 weeks gestation | Beta | 0.019 | 0.001, 0.036 | 0.036 | 0.015, 0.036 | 0.005 | 6742 |
| Sleep problems 32 weeks gestation | Beta | 0.028 | 0.009, 0.046 | 0.003 | <0.001, 0.004 | <0.001 | 6743 |
| Total number of outcomes tested = 22 | | | | | | | |
| **Offspring: Adolescents** | | | | | | | |
| **Mental health** | | | | | | | |
| Conduct disorder symptoms | Beta | 0.041 | 0.015, 0.066 | 0.002 | <0.001, 0.006 | 0.001 | 3834 |
| ADHD symptoms | Beta | 0.050 | 0.016, 0.085 | 0.004 | 0.001, 0.010 | 0.004 | 3852 |
| Oppositional-defiant disorder symptoms | Beta | 0.036 | 0.011, 0.060 | 0.004 | <0.001, 0.006 | 0.001 | 3436 |
| Psychosis positive symptoms age 12 | Beta | 0.015 | <0.001, 0.029 | 0.046 | 0.034, 0.061 | 0.046 | 4974 |
| Psychosis negative symptoms age 16 | Beta | -0.022 | -0.059, 0.015 | 0.251 | 0.230, 0.285 | 0.257 | 3511 |
| Psychosis positive symptoms age 18 | Beta | 0.012 | -0.004, 0.028 | 0.134 | 0.089, 0.129 | 0.134 | 3403 |
| PTSD disorder | Beta | 0.013 | -0.002, 0.028 | 0.085 | 0.042, 0.071 | 0.055 | 4008 |
| Depression score age 17 (MFQ) | Beta | 0.008 | -0.004, 0.020 | 0.178 | 0.164, 0.214 | 0.188 | 3212 |
| Depression symptom score age 18 (CIS-R) | Beta | 0.015 | -0.010, 0.041 | 0.236 | 0.210, 0.264 | 0.236 | 3303 |
| Eating disorder age 16 | Beta | -0.002 | -0.007, 0.002 | 0.281 | 0.247, 0.303 | 0.274 | 3543 |
| Eating disorder age 13 | Beta | -0.001 | -0.003, 0.001 | 0.395 | 0.452, 0.514 | 0.483 | 4256 |
| Specific phobia symptoms | Beta | 0.010 | -0.008, 0.028 | 0.298 | 0.269,0.326 | 0.297 | 3293 |
| Emotional problems symptoms | Beta | -0.009 | -0.032, 0.014 | 0.422 | 0.410, 0.472 | 0.441 | 4073 |
| Self-harming behaviour | OR | 0.958 | 0.810, 1.135 | 0.596 | 0.562, 0.624 | 0.593 | 2576 |
| Depression symptoms score age 14 (MFQ) | Beta | -0.002 | -0.016, 0.013 | 0.821 | 0.808, 0.856 | 0.833 | 4574 |
| Anxiety score | Beta | 0.002 | -0.023, 0.027 | 0.848 | 0.830, 0.874 | 0.853 | 3293 |
| Total behavioural difficulties score | Beta | 0.036 | -0.001, 0.073 | 0.055 | 0.035, 0.062 | 0.047 | 4055 |
| **Substance use** | | | | | | | |
| Cannabis use | OR | 1.225 | 1.127, 1.330 | <0.001 | <0.001, 0.004 | <0.001 | 3571 |
| AUDIT risk score age 18 | Beta | 0.041 | 0.018, 0.064 | 0.001 | <0.001, 0.004 | <0.001 | 3008 |
| Binge drinking age 18 | Beta | 0.070 | 0.025, 0.114 | 0.002 | 0.001, 0.010 | 0.004 | 2829 |
| AUDIT total score age 18 | Beta | 0.069 | 0.029, 0.109 | 6.58x10^-4^ | <0.001, 0.004 | <0.001 | 3008 |
| Number of alcoholic drinks on a typical day | Beta | 0.066 | 0.022, 0.111 | 0.003 | <0.001, 0.007 | 0.002 | 2826 |
| Number of drinks to feel tipsy | Beta | 0.050 | 0.008, 0.093 | 0.020 | 0.017, 0.038 | 0.026 | 2391 |
| Number of drinks to feel different after first five times drinking | Beta | 0.097 | -0.016, 0.211 | 0.093 | 0.073, 0.109 | 0.090 | 299 |
| Binge drinking age 13 | Beta | 0.039 | -0.065, 0.142 | 0.461 | 0.421, 0.483 | 0.452 | 464 |
| Frequency of having alcoholic drinks | Beta | 0.008 | -0.024, 0.039 | 0.641 | 0.586, 0.647 | 0.617 | 3626 |
| Number of times had whole drink age 13 | Beta | 0.007 | -0.066, 0.079 | 0.860 | 0.838, 0.882 | 0.861 | 1103 |
| Frequency of cannabis smoking | Beta | -0.016 | -0.092, 0.060 | 0.676 | 0.625, 0.684 | 0.655 | 1035 |
| Total caffeine age 13 | Beta | 0.008 | -0.030, 0.046 | 0.680 | 0.639, 0.698 | 0.669 | 3405 |
| **Non-mental health** | | | | | | | |
| BMI | Beta | 0.239 | 0.104, 0.373 | 0.001 | <0.001, 0.004 | <0.001 | 3606 |
| IQ | Beta | -0.582 | -1.006, -0.159 | 0.007 | 0.003, 0.016 | 0.008 | 3720 |
| GCSE grades D-G | OR | 1.086 | 0.988, 1.193 | 0.083 | 0.040, 0.069 | 0.053 | 2182 |
| GCSE grades A-C | OR | 0.819 | 0.651, 1.032 | 0.085 | 0.063, 0.097 | 0.079 | 2360 |
| Extraversion personality trait | Beta | 0.362 | 0.155, 0.569 | 0.001 | <0.001, 0.006 | 0.001 | 4354 |
| Conscientiousness personality trait | Beta | -0.217 | -0.403, -0.031 | 0.022 | 0.006, 0.021 | 0.012 | 4162 |
| Emotional Stability personality trait | Beta | -0.073 | -0.263, 0.117 | 0.449 | 0.436, 0.498 | 0.467 | 4224 |
| Intellect personality trait | Beta | -0.053 | -0.226, 0.119 | 0.545 | 0.491, 0.553 | 0.522 | 4263 |
| Agreeableness personality trait | Beta | -0.037 | -0.188, 0.113 | 0.628 | 0.610, 0.671 | 0.641 | 4279 |
| Sleep maintenance | Beta | 0.025 | <0.001, 0.051 | 0.051 | 0.039, 0.068 | 0.052 | 3418 |
| Sleep initiation (time to fall asleep) | Beta | 0.008 | -0.027, 0.043 | 0.641 | 0.616, 0.677 | 0.647 | 3626 |
| Sleep duration (hours of sleep) | Beta | 0.015 | -0.017, 0.047 | 0.360 | 0.304, 0.363 | 0.333 | 3726 |
| Frequency of doing exercise | Beta | -0.004 | -0.027, 0.020 | 0.762 | 0.736, 0.790 | 0.764 | 4270 |
| Life events | Beta | -0.006 | -0.043, 0.031 | 0.756 | 0.762, 0.814 | 0.789 | 3376 |
| Total number of outcomes tested = 44 | | | | | | | |

**Supplementary table S6. Associations between maternal and offspring lifetime smoking GRS and phenotypes in mothers during and outside pregnancy and adolescence**

|  |  | **Regression analyses** | | | **Permutation testing** | | |
| --- | --- | --- | --- | --- | --- | --- | --- |
| **Phenotype** | **Effect estimate** | **Effect size** | **95% CI** | **P-value** | **95% CI** | **P-value** | **Sample size** |
| **Mothers outside of pregnancy** | | | | | | | |
| **Mental health** | | | | | | | |
| Depression symptoms | OR | 1.007 | 0.913, 1.109 | 0.886 | 0.876, 0.915 | 0.897 | 4725 |
| Anxiety symptoms | OR | 1.005 | 0.915, 1.105 | 0.904 | 0.880, 0.918 | 0.900 | 4740 |
| Bulimia | OR | 1.140 | 0.964, 1.349 | 0.114 | 0.083, 0.121 | 0.101 | 6799 |
| Drug addiction | OR | 0.983 | 0.596, 1.620 | 0.941 | 0.912, 0.945 | 0.930 | 6799 |
| Alcoholism | OR | 1.270 | 0.968, 1.667 | 0.079 | 0.101, 0.143 | 0.121 | 6799 |
| Schizophrenia | OR | 1.585 | 0.870, 2.889 | 0.120 | 0.197, 0.249 | 0.222 | 6799 |
| Anorexia Nervosa | OR | 1.150 | 0.932, 1.419 | 0.174 | 0.096, 0.136 | 0.115 | 6799 |
| Severe depression | OR | 1.159 | 1.049, 1.280 | 0.007 | 0.001, 0.010 | 0.004 | 6799 |
| Other psychiatric problem | OR | 1.156 | 0.949, 1.408 | 0.134 | 0.079, 0.117 | 0.097 | 6799 |
| **Substance use** | | | | | | | |
| *Alcohol* | | | | | | | |
| Alcohol drinking before pregnancy | OR | 1.010 | 0.910, 1.122 | 0.833 | 0.816, 0.862 | 0.840 | 7199 |
| Binge drinking | Beta | 0.039 | 0.009, 0.068 | 0.010 | <0.001, 0.004 | <0.001 | 4867 |
| Daily alcohol units at child age 4 | Beta | 0.028 | 0.007, 0.049 | 0.008 | 0.006, 0.020 | 0.011 | 5680 |
| Daily alcohol units at child age 8 | Beta | -0.009 | -0.039, 0.021 | 0.559 | 0.564, 0.626 | 0.595 | 2707 |
| AUDIT score | Beta | 0.014 | -0.007, 0.035 | 0.181 | 0.174, 0.224 | 0.198 | 2424 |
| *Caffeine* | | | | | | | |
| Total caffeine consumption | Beta | 8.698 | 5.083, 12.313 | 2.46 x 10^-6^ | <0.001, 0.004 | <0.001 | 4783 |
| **Non-mental health** | | | | | | | |
| Life events | Beta | 0.026 | -0.009, 0.060 | 0.142 | 0.117, 0.161 | 0.138 | 4219 |
| Sleep duration | Beta | -0.018 | -0.046, 0.009 | 0.183 | 0.163, 0.213 | 0.187 | 1867 |
| Impulsivity personality trait | Beta | 0.111 | 0.006, 0.217 | 0.039 | 0.033, 0.060 | 0.045 | 4847 |
| Monotony avoidance personality trait | Beta | 0.184 | 0.037, 0.332 | 0.014 | 0.008, 0.025 | 0.015 | 4794 |
| Anger personality trait | Beta | 0.246 | 0.115, 0.377 | 2.34x10^-4^ | <0.001, 0.004 | <0.001 | 4769 |
| Suspicion personality trait | Beta | 0.164 | 0.057, 0.272 | 0.003 | 0.003, 0.016 | 0.008 | 4856 |
| Detachment personality trait | Beta | -0.061 | -0.175, 0.054 | 0.301 | 0.258, 0.315 | 0.286 | 4753 |
| Physical activity | OR | 0.996 | 0.915, 1.085 | 0.929 | 0.919, 0.950 | 0.936 | 2787 |
| Social class | Beta | 0.028 | -0.015, 0.072 | 0.204 | 0.185, 0.237 | 0.210 | 2906 |
| Education | Beta | -0.083 | -0.115, -0.051 | 4.33 x 10^-7^ | <0.001, 0.004 | <0.001 | 4919 |
| BMI before pregnancy | Beta | 0.160 | 0.065, 0.254 | 0.001 | 0.001, 0.010 | 0.004 | 6398 |
| Image perception before pregnancy | Beta | 0.028 | 0.002, 0.054 | 0.037 | 0.029, 0.054 | 0.040 | 6623 |
| Total number of outcomes tested = 27 |  |  |  |  |  |  |  |
| **Mothers during pregnancy** | | | | | | | |
| **Mental health** | | | | | | | |
| Depression (18wks) | OR | 1.076 | 0.997, 1.163 | 0.060 | 0.034, 0.061 | 0.046 | 6734 |
| Depression (32wks) | OR | 1.078 | 0.999, 1.164 | 0.053 | 0.015, 0.036 | 0.024 | 6751 |
| Anxiety (18 wks) | OR | 1.064 | 0.980, 1.155 | 0.127 | 0.087, 0.126 | 0.105 | 6645 |
| Hypersensitivity to interpersonal rejection | Beta | -0.296 | -0.657, 0.065 | 0.108 | 0.097, 0.137 | 0.116 | 7167 |
| Feelings becoming a parent | Beta | -0.012 | -0.034, 0.009 | 0.266 | 0.236, 0.291 | 0.263 | 7165 |
| **Substance use** | | | | | | | |
| *Caffeine* | | | | | | | |
| Total caffeine (18wks) | Beta | 6.759 | 4.239, 9.280 | <0.001 | <0.001, 0.004 | <0.001 | 7220 |
| Total caffeine (32wks) | Beta | 5.325 | 2.776, 7.874 | <0.001 | <0.001, 0.004 | <0.001 | 6767 |
| *Alcohol* | | | | | | | |
| Binge drinking (18wks) | Beta | 0.024 | 0.005, 0.042 | 0.012 | 0.003, 0.016 | 0.008 | 7171 |
| Binge drinking (32wks) | Beta | 0.020 | 0.001, 0.039 | 0.044 | 0.049, 0.080 | 0.063 | 5324 |
| Weekly alcohol units (32wks) | Beta | 0.133 | 0.034, 0.233 | 0.009 | 0.008, 0.025 | 0.015 | 4294 |
| *Other substances* | | | | | | | |
| Cannabis use during pregnancy | OR | 1.106 | 0.942, 1.299 | 0.197 | 0.175, 0.225 | 0.199 | 6918 |
| Hard drugs | OR | 1.053 | 0.670, 1.653 | 0.809 | 0.779, 0.829 | 0.805 | 7147 |
| **Non-mental health** | | | | | | | |
| Education | Beta | -0.094 | -0.122, -0.065 | <0.001 | <0.001, 0.004 | <0.001 | 6954 |
| Social class | Beta | 0.064 | 0.037, 0.091 | <0.001 | <0.001, 0.004 | <0.001 | 5854 |
| Life events during pregnancy | Beta | 0.018 | -0.010, 0.045 | 0.214 | 0.196, 0.248 | 0.221 | 6744 |
| Image perception during pregnancy | Beta | 0.121 | 0.023, 0.219 | 0.016 | 0.011, 0.028 | 0.018 | 6699 |
| Image perception change | Beta | 0.004 | -0.087, 0.095 | 0.931 | 0.906, 0.940 | 0.924 | 6549 |
| Activity level compared with other pregnant women | Beta | -0.001 | -0.019, 0.017 | 0.911 | 0.892, 0.928 | 0.911 | 6611 |
| Physical activity | OR | 1.002 | 0.946, 1.061 | 0.952 | 0.941, 0.968 | 0.956 | 6767 |
| Vomited first three months in pregnancy | OR | 0.979 | 0.928, 1.033 | 0.412 | 0.373, 0.435 | 0.404 | 6797 |
| Sleep (18 wks) | Beta | 0.014 | -0.003, 0.032 | 0.108 | 0.089, 0.129 | 0.108 | 6742 |
| Sleep (32 wks) | Beta | 0.034 | 0.016, 0.052 | <0.001 | <0.001, 0.004 | <0.001 | 6743 |
| **Offspring: Adolescence** | | | | | | | |
| **Mental health** | | | | | | | |
| Conduct disorder symptoms | Beta | 0.057 | 0.031, 0.082 | <0.001 | <0.001, 0.004 | <0.001 | 3834 |
| Psychosis positive symptoms age 12 | Beta | 0.024 | 0.010, 0.018 | 0.001 | <0.001, 0.004 | <0.001 | 4974 |
| Depression symptoms age 17 (MFQ) | Beta | 0.019 | 0.007, 0.036 | 0.002 | <0.001, 0.006 | 0.001 | 3212 |
| Total behavioural difficulties | Beta | 0.055 | 0.019, 0.091 | 0.003 | 0.001, 0.009 | 0.003 | 4055 |
| Psychosis positive symptoms age 18 | Beta | 0.020 | 0.004, 0.018 | 0.014 | 0.004, 0.017 | 0.009 | 3403 |
| ADHD symptoms | Beta | 0.031 | -0.003, 0.030 | 0.075 | 0.078, 0.116 | 0.096 | 3852 |
| Eating disorder age 16 | Beta | 0.003 | -0.001, 0.041 | 0.146 | 0.138, 0.184 | 0.160 | 3543 |
| Depression symptoms score age 17 | Beta | 0.016 | -0.010, 0.039 | 0.228 | 0.218, 0.272 | 0.244 | 3303 |
| Specific phobia symptoms | Beta | 0.010 | -0.009, 0.031 | 0.301 | 0.261, 0.318 | 0.289 | 3293 |
| PTSD symptoms | Beta | 0.006 | -0.007, 0.027 | 0.360 | 0.340, 0.401 | 0.370 | 4008 |
| Oppositional defiant disorder | Beta | 0.011 | -0.013, 0.169 | 0.374 | 0.342, 0.403 | 0.372 | 3436 |
| Anxiety symptoms score | Beta | 0.012 | -0.015, 0.066 | 0.387 | 0.345, 0.406 | 0.375 | 3293 |
| Eating disorder age 13 | Beta | 0.001 | -0.001, 0.007 | 0.475 | 0.454, 0.516 | 0.485 | 4256 |
| Depression symptoms age 14 (MFQ) | Beta | 0.004 | -0.010, 0.019 | 0.573 | 0.568, 0.630 | 0.599 | 4574 |
| Emotional problems symptoms | Beta | 0.004 | -0.018, 0.036 | 0.700 | 0.655, 0.714 | 0.685 | 4073 |
| Psychosis negative symptoms age 16 | Beta | 0.002 | -0.034, 0.038 | 0.922 | 0.892, 0.928 | 0.911 | 3511 |
| Self-harming behaviour | OR | 0.994 | 0.834, 1.185 | 0.944 | 0.927, 0.957 | 0.943 | 2576 |
| **Substance use** | | | | | | | |
| *Alcohol* | | | | | | | |
| Number of drinks needed to feel different | Beta | 0.099 | -0.020, 0.218 | 0.103 | 0.101, 0.143 | 0.121 | 299 |
| Binge drinking age 13 | Beta | 0.090 | -0.017, 0.197 | 0.099 | 0.073, 0.109 | 0.090 | 464 |
| Number of times had a whole drink past 6 months | Beta | -0.007 | -0.076, 0.062 | 0.840 | 0.825, 0.871 | 0.849 | 1103 |
| Number of alcoholic drinks on a typical day | Beta | 0.039 | -0.005, 0.082 | 0.083 | 0.058, 0.091 | 0.073 | 2826 |
| Binge drinking age 18 | Beta | 0.063 | 0.017, 0.109 | 0.007 | 0.002, 0.012 | 0.005 | 2829 |
| Frequency of having alcoholic drinks | Beta | -0.002 | -0.032, 0.027 | 0.876 | 0.833, 0.877 | 0.856 | 2886 |
| AUDIT risk score age 18 | Beta | 0.038 | 0.014, 0.061 | 0.002 | <0.001, 0.006 | 0.001 | 3008 |
| AUDIT total score age 18 | Beta | 0.035 | -0.004, 0.075 | 0.082 | 0.088, 0.127 | 0.106 | 3008 |
| Number of drinks needed to feel tipsy | Beta | 0.047 | 0.004, 0.089 | 0.032 | 0.030, 0.055 | 0.041 | 2391 |
| *Tobacco* | | | | | | | |
| Cannabis use | OR | 1.074 | 0.990, 1.164 | 0.082 | 0.040, 0.069 | 0.053 | 3571 |
| Frequency of cannabis use | Beta | 0.042 | -0.031, 0.115 | 0.261 | 0.248, 0.305 | 0.276 | 1035 |
| *Caffeine* | | | | | | | |
| Total caffeine consumption | Beta | 0.018 | -0.019, 0.055 | 0.348 | 0.342, 0.403 | 0.372 | 3405 |
| **Non-mental health** | | | | | | | |
| Extraversion personality trait | Beta | 0.445 | 0.244, 0.646 | <0.001 | <0.001, 0.004 | <0.001 | 4354 |
| Conscientiousness personality trait | Beta | -0.187 | -0.367, -0.008 | 0.041 | 0.030, 0.056 | 0.042 | 4162 |
| Agreeableness personality trait | Beta | 0.010 | -0.132, 0.151 | 0.893 | 0.896, 0.932 | 0.915 | 4279 |
| Intellect personality trait | Beta | -0.009 | -0.174, 0.156 | 0.918 | 0.900, 0.935 | 0.919 | 4263 |
| Emotional stability personality trait | Beta | -0.065 | -0.253, 0.124 | 0.501 | 0.489, 0.551 | 0.520 | 4224 |
| IQ | Beta | -0.741 | -1.163, -0.320 | 0.001 | <0.001, 0.004 | <0.001 | 3720 |
| BMI | Beta | 0.205 | 0.078, 0.331 | 0.001 | 0.001, 0.010 | 0.004 | 3606 |
| Sleep maintenance | Beta | 0.033 | 0.008, 0.059 | 0.011 | 0.005, 0.018 | 0.010 | 3418 |
| GCSE grades D-G | OR | 1.106 | 1.008, 1.213 | 0.036 | 0.011, 0.028 | 0.018 | 2182 |
| Frequency of doing exercise | Beta | -0.025 | -0.048, -0.001 | 0.039 | 0.032, 0.059 | 0.044 | 4270 |
| Sleep duration (hours of sleep) | Beta | -0.025 | -0.055, 0.005 | 0.097 | 0.088, 0.127 | 0.106 | 3726 |
| Sleep initiation (time to fall asleep) | Beta | 0.025 | -0.009, 0.060 | 0.150 | 0.127, 0.173 | 0.149 | 3626 |
| GCSE grades A-C | OR | 0.840 | 0.652, 1.082 | 0.160 | 0.101, 0.142 | 0.120 | 2360 |
| Life events | Beta | 0.011 | -0.026, 0.047 | 0.570 | 0.583, 0.644 | 0.614 | 3376 |
| Total number of outcomes tested = 22 |  |  |  |  |  |  |  |

**Supplementary table S7. Associations between caffeine GRS and phenotypes in mothers during and outside of pregnancy and adolescence**

|  |  | **Regression analyses** | | | **Permutation testing** | | | |
| --- | --- | --- | --- | --- | --- | --- | --- | --- |
| **Phenotype** | **Effect estimate** | **Effect size** | **95% CI** | **P-value** | **95% CI** | **P-value** | | **Sample size** |
| **Mothers outside of pregnancy** | | | | | | | | |
| **Mental health** | | | | | | | | |
| Depression symptoms | OR | 1.040 | 0.943, 1.148 | 0.398 | 0.507, 0.569 | 0.538 | | 4725 |
| Anxiety symptoms | OR | 0.979 | 0.890, 1.077 | 0.641 | 0.340, 0.401 | 0.370 | | 4740 |
| Bulimia | OR | 1.105 | 0.932, 1.309 | 0.225 | 0.180, 0.231 | 0.205 | | 6799 |
| Drug addiction | OR | 0.987 | 0.657, 1.481 | 0.943 | 0.941, 0.968 | 0.956 | | 6799 |
| Alcoholism | OR | 0.947 | 0.697, 1.287 | 0.706 | 0.697, 0.753 | 0.726 | | 6799 |
| Schizophrenia | OR | 0.434 | 0.244, 0.772 | 0.008 | 0.021, 0.044 | 0.031 | | 6799 |
| Anorexia Nervosa | OR | 1.076 | 0.899, 1.289 | 0.390 | 0.354, 0.415 | 0.384 | | 6799 |
| Severe depression | OR | 1.054 | 0.953, 1.166 | 0.277 | 0.218, 0.272 | 0.244 | | 6799 |
| Other psychiatric problem | OR | 1.048 | 0.867, 1.266 | 0.601 | 0.529, 0.591 | 0.560 | | 6799 |
| **Substance use** | | | | | | | | |
| *Tobacco* | | | | | | | | |
| Ever smoking | OR | 1.010 | 0.959, 1.064 | 0.679 | 0.626. 0.685 | 0.656 | | 7194 |
| Number of cigarettes smoked past 2 weeks | Beta | 0.304 | -0.271, 0.879 | 0.300 | 0.270, 0.327 | 0.298 | | 845 |
| Number of cigarettes smoked before pregnancy | Beta | 0.042 | -0.028, 0.111 | 0.245 | 0.224, 0.279 | 0.251 | | 3426 |
| *Alcohol* | | | | | | | | |
| Alcohol drinking before pregnancy | OR | 0.972 | 0.876, 1.078 | 0.558 | 0.016, 0.514 | 0.545 | | 7199 |
| Binge drinking | Beta | 0.004 | -0.024, 0.032 | 0.786 | 0.752, 0.804 | 0.779 | | 4867 |
| Daily alcohol units at child age 4 | Beta | 0.007 | -0.013, 0.027 | 0.518 | 0.467, 0.529 | 0.498 | | 5680 |
| Daily alcohol units at child age 8 | Beta | 0.014 | -0.016, 0.044 | 0.347 | 0.354, 0.415 | 0.384 | | 2707 |
| AUDIT score | Beta | 0.007 | -0.013, 0.028 | 0.473 | 0.450, 0.512 | 0.481 | | 2424 |
| **Non-mental health** | | | | | | | | |
| Life events | Beta | -0.025 | -0.059, 0.008 | 0.141 | 0.115, 0.159 | 0.136 | | 4219 |
| Sleep duration | Beta | -0.007 | -0.034, 0.019 | 0.588 | 0.535, 0.597 | 0.566 | | 1867 |
| Impulsivity personality trait | Beta | 0.042 | -0.063, 0.146 | 0.436 | 0.397, 0.459 | 0.428 | | 4847 |
| Monotony avoidance personality trait | Beta | -0.107 | -0.251, 0.037 | 0.144 | 0.131, 0.177 | 0.153 | | 4794 |
| Anger personality trait | Beta | 0.014 | -0.115, 0.144 | 0.830 | 0.784, 0.834 | 0.810 | | 4769 |
| Suspicion personality trait | Beta | -0.017 | -0.128, 0.095 | 0.772 | 0.718, 0.773 | 0.746 | | 4856 |
| Detachment personality trait | Beta | 0.012 | -0.102, 0.125 | 0.841 | 0.815, 0.861 | 0.839 | | 4753 |
| Physical activity | OR | 0.966 | 0.890, 1.050 | 0.387 | 0.015, 0.340 | 0.370 | | 2787 |
| Social class | Beta | -0.009 | -0.053, 0.035 | 0.696 | 0.680, 0.737 | 0.709 | | 2906 |
| Education | Beta | 0.002 | -0.030, 0.035 | 0.889 | 0.882, 0.920 | 0.902 | | 4919 |
| BMI before pregnancy | Beta | 0.083 | -0.008, 0.174 | 0.075 | 0.058, 0.091 | 0.073 | | 6398 |
| Image perception before pregnancy | Beta | -0.003 | -0.029, 0.023 | 0.820 | 0.796, 0.844 | 0.821 | | 6623 |
| Total number of outcomes tested = 29 |  |  |  |  |  |  | |  |
| **Mothers during pregnancy** | | | | | | | | |
| **Mental health** | | | | | | | | |
| Depression symptoms (18 wks) | OR | 0.982 | 0.905, 1.067 | 0.647 | 0.616, 0.677 | 0.647 | | 6734 |
| Depression symptoms (32 wks) | OR | 0.994 | 0.920, 1.074 | 0.870 | 0.823, 0.869 | 0.847 | | 6751 |
| Anxiety symptoms | OR | 1.005 | 0.919, 1.099 | 0.910 | 0.891, 0.927 | 0.910 | | 6645 |
| Hypersensitivity to interpersonal rejection | Beta | -0.040 | 0.838, 0.882 | 0.833 | 0.838, 0.882 | 0.861 | | 7167 |
| Feelings becoming a parent | Beta | 0.007 | 0.496, 0.558 | 0.528 | 0.496, 0.558 | 0.527 | | 7165 |
| **Substance use** | | | | | | | | |
| *Tobacco* | | | | | | | | |
| Ever smoked in pregnancy | OR | 1.007 | 0.947, 1.070 | 0.813 | 0.783, 0.833 | 0.809 | | 6718 |
| Smoking first three months in pregnancy | OR | 1.029 | 0.966, 1.097 | 0.343 | 0.275, 0.333 | 0.303 | | 7237 |
| *Caffeine* | | | | | | | | |
| Reduced caffeine consumption during pregnancy | OR | 1.054 | 1.001, 1.111 | 0.046 | 0.017, 0.038 | 0.026 | | 7269 |
| Reduced coffee consumption during pregnancy | OR | 1.061 | 1.008, 1.117 | 0.028 | 0.006, 0.021 | 0.012 | | 7269 |
| Stopped drinking cola during pregnancy | OR | 1.094 | 0.991, 1.208 | 0.072 | 0.034, 0.061 | 0.046 | | 4570 |
| Never drank coffee | OR | 1.064 | 0.988, 1.146 | 0.093 | 0.059, 0.092 | 0.074 | | 6782 |
| Never drank cola | OR | 1.002 | 0.946, 1.062 | 0.933 | 0.915, 0.947 | 0.932 | | 6744 |
| Stopped drinking coffee during pregnancy | OR | 1.039 | 0.981, 1.100 | 0.175 | 0.104, 0.146 | 0.124 | | 5809 |
| Never has been drinking caffeine | OR | 0.967 | 0.918, 1.018 | 0.179 | 0.126, 0.170 | 0.147 | | 7269 |
| Stopped drinking tea during pregnancy | OR | 1.039 | 0.973, 1.109 | 0.226 | 0.184, 0.236 | 0.209 | | 6082 |
| Reduced cola consumption during pregnancy | OR | 1.039 | 0.967, 1.116 | 0.272 | 0.244, 0.300 | 0.271 | | 7269 |
| Never drank tea | OR | 1.041 | 0.950, 1.141 | 0.353 | 0.269, 0.326 | 0.297 | | 6754 |
| Reduced tea consumption during pregnancy | OR | 1.026 | 0.970, 1.085 | 0.333 | 0.290, 0.349 | 0.319 | | 7269 |
| Consumed more caffeine during pregnancy | OR | 1.038 | 0.948, 1.137 | 0.384 | 0.333, 0.394 | 0.363 | | 7269 |
| No change in caffeine consumption during pregnancy | OR | 0.986 | 0.934, 1.041 | 0.580 | 0.532, 0.594 | 0.563 | | 7269 |
| Craved or had more caffeine during pregnancy | OR | 0.985 | 0.909, 1.068 | 0.698 | 0.682, 0.739 | 0.711 | | 7269 |
| Craved or had more coffee during pregnancy | OR | 0.977 | 0.824, 1.160 | 0.774 | 0.700, 0.756 | 0.729 | | 6782 |
| Craved or had more tea during pregnancy | OR | 1.013 | 0.927, 1.108 | 0.750 | 0.718, 0.773 | 0.746 | | 6754 |
| *Alcohol* | | | | | | | | |
| Binge drinking (32wks) | Beta | -0.022 | -0.041, -0.003 | 0.026 | 0.031, 0.057 | 0.043 | | 5324 |
| Binge drinking (18wks) | Beta | -0.010 | -0.029, 0.008 | 0.268 | 0.249, 0.306 | 0.277 | | 7171 |
| Weekly alcohol units (32wks) | Beta | -0.056 | -0.167, 0.056 | 0.329 | 0.314, 0.373 | 0.343 | | 4294 |
| Craved or had more alcohol during pregnancy | OR | 0.949 | 0.569, 1.584 | 0.828 | 0.826, 0.872 | 0.850 | | 6771 |
| *Other substances* | | | | | | | | |
| Cannabis use in first three months during pregnancy | OR | 1.124 | 0.952, 1.328 | 0.151 | 0.134, 0.180 | 0.156 | | 6918 |
| Hard drugs during pregnancy | OR | 0.995 | 0.664, 1.491 | 0.980 | 0.968, 0.987 | 0.979 | | 7147 |
| **Non-mental health** | | | | | | | | |
| Life events during pregnancy | Beta | 0.001 | 0.154, 0.202 | 0.944 | 0.154, 0.202 | 0.177 | | 6930 |
| Activity level compared with other pregnant women | Beta | 0.011 | 0.227, 0.282 | 0.234 | 0.227, 0.282 | 0.254 | | 6611 |
| Image perception during pregnancy | Beta | -0.031 | 0.517, 0.579 | 0.540 | 0.517, 0.579 | 0.548 | | 6699 |
| Physical activity | Beta | -0.003 | 0.754, 0.806 | 0.780 | 0.754, 0.806 | 0.781 | | 6767 |
| Social class | Beta | 0.028 | 0.035, 0.062 | 0.043 | 0.035, 0.062 | 0.047 | | 6954 |
| Image perception change | Beta | 0.019 | 0.394, 0.456 | 0.678 | 0.394, 0.456 | 0.425 | | 3741 |
| Education | Beta | -0.005 | 0.658, 0.717 | 0.709 | 0.658, 0.717 | 0.688 | | 6954 |
| Vomiting in first three months during pregnancy | OR | 1.003 | 0.950, 1.059 | 0.903 | 0.871, 0.911 | 0.892 | | 0.903 |
| Sleeping problems (18 wks) | Beta | 0.002 | 0.797, 0.845 | 0.825 | 0.797, 0.845 | 0.822 | | 6742 |
| Sleeping problems (32 wks) | Beta | -0.003 | 0.726, 0.780 | 0.733 | 0.726, 0.780 | 0.754 | | 6743 |
| Total number of outcomes tested = 39 |  |  |  |  |  |  | |  |
| **Offspring: Adolescence** | | | | | | | | |
| **Mental health** | | | | | | | | |
| Conduct disorder symptoms | Beta | 0.012 | -0.014, 0.039 | 0.362 | 0.332, 0.393 | 0.362 | 3834 | |
| Depression symptoms score age 18 | Beta | 0.013 | -0.013, 0.039 | 0.314 | 0.288, 0.347 | 0.317 | 3303 | |
| Specific phobia symptoms | Beta | 0.001 | -0.019, 0.020 | 0.937 | 0.935, 0.963 | 0.950 | 3293 | |
| Emotional problems score | Beta | -0.022 | -0.047, 0.002 | 0.072 | 0.063, 0.097 | 0.079 | 3593 | |
| Anxiety symptoms | Beta | 0.001 | -0.024, 0.026 | 0.913 | 0.899, 0.934 | 0.918 | 3293 | |
| Eating disorder age 13 | Beta | -0.001 | -0.003, 0.001 | 0.289 | 0.346, 0.407 | 0.376 | 4256 | |
| Eating disorder age 16 | Beta | 0.003 | -0.001, 0.007 | 0.184 | 0.166, 0.216 | 0.190 | 3543 | |
| ADHD symptoms | Beta | -0.027 | -0.065, 0.010 | 0.146 | 0.142, 0.188 | 0.164 | 3435 | |
| Depression score age 14 (MFQ) | Beta | -0.002 | -0.016, 0.013 | 0.835 | 0.817, 0.863 | 0.841 | 4574 | |
| Depression score age 17 (MFQ) | Beta | -0.003 | -0.015, 0.010 | 0.685 | 0.647, 0.706 | 0.677 | 3212 | |
| Psychosis negative symptoms age 16 | Beta | 0.000 | -0.037, 0.036 | 0.996 | 0.993, 1.000 | 0.998 | 3511 | |
| Total behavioural difficulties | Beta | -0.017 | -0.056, 0.022 | 0.397 | 0.364, 0.425 | 0.394 | 3603 | |
| Psychosis positive symptoms age 12 | Beta | 0.009 | -0.006, 0.024 | 0.230 | 0.198, 0.250 | 0.223 | 4974 | |
| Psychosis positive symptoms age 18 | Beta | 0.011 | -0.006, 0.027 | 0.200 | 0.097, 0.137 | 0.116 | 3403 | |
| PTSD disorder symptoms | Beta | -0.013 | -0.027, 0.002 | 0.091 | 0.052, 0.084 | 0.067 | 4008 | |
| Self-harming behaviour | OR | 0.985 | 0.811, 1.196 | 0.869 | 0.836, 0.880 | 0.859 | 2576 | |
| Oppositional-defiant disorder symptoms age 15 | Beta | -0.011 | -0.036, 0.013 | 0.367 | 0.561, 0.623 | 0.592 | 3436 | |
| **Substance use** | | | | | | | | |
| *Tobacco* | | | | | | | | |
| Age when first smoked a cigarette | Beta | -0.013 | -0.056, 0.030 | 0.553 | 0.535, 0.597 | 0.566 | 1064 | |
| Has smoked a cigarette | OR | 1.045 | 0.927, 1.179 | 0.443 | 0.384, 0.446 | 0.415 | 2089 | |
| Total number of cigarettes smoked age 14 | OR | 0.990 | 0.767, 1.277 | 0.931 | 0.900, 0.935 | 0.919 | 461 | |
| Total number of cigarettes smoked age 18 | Beta | 0.069 | -0.023, 0.162 | 0.142 | 0.114, 0.158 | 0.135 | 1144 | |
| *Alcohol* |  |  |  |  |  |  |  | |
| Number of drinks to feel different | Beta | -0.039 | -0.154, 0.076 | 0.505 | 0.494, 0.556 | 0.525 | 299 | |
| Binge drinking age 13 | Beta | 0.010 | -0.094, 0.113 | 0.854 | 0.834, 0.878 | 0.857 | 464 | |
| Number of times had whole drink age 13 | Beta | 0.012 | -0.059, 0.083 | 0.748 | 0.700, 0.756 | 0.729 | 1103 | |
| Number of alcoholic drinks on a typical day | Beta | -0.012 | -0.058, 0.034 | 0.609 | 0.580, 0.641 | 0.611 | 2826 | |
| Binge drinking age 18 | Beta | 0.010 | -0.036, 0.056 | 0.670 | 0.632, 0.691 | 0.662 | 2829 | |
| Frequency having alcoholic drinks | Beta | 0.011 | -0.020, 0.042 | 0.485 | 0.462, 0.524 | 0.493 | 2886 | |
| AUDIT risk score age 18 | Beta | -0.007 | -0.030, 0.017 | 0.562 | 0.539, 0.601 | 0.570 | 3008 | |
| AUDIT total score age 18 | Beta | 0.010 | -0.031, 0.050 | 0.647 | 0.986, 0.997 | 0.993 | 3008 | |
| Number of drinks needed to feel tipsy | Beta | -0.015 | -0.059, 0.029 | 0.500 | 0.461, 0.523 | 0.492 | 2391 | |
| *Other substances* |  |  |  |  |  |  |  | |
| Cannabis use | OR | 0.977 | 0.900, 1.060 | 0.551 | 0.494, 0.556 | 0.525 | 3571 | |
| Frequency of cannabis use | Beta | 0.018 | -0.057, 0.093 | 0.636 | 0.613, 0.674 | 0.644 | 1035 | |
| **Non-mental health** | | | | | | | | |
| BMI | Beta | 0.031 | -0.100, 0.161 | 0.645 | 0.612, 0.673 | 0.643 | 3606 | |
| Agreeableness personality trait | Beta | 0.066 | -0.080, 0.211 | 0.376 | 0.368, 0.430 | 0.399 | 4279 | |
| Conscientiousness personality trait | Beta | -0.044 | -0.218, 0.130 | 0.617 | 0.600, 0.661 | 0.631 | 4162 | |
| Intellect personality trait | Beta | 0.100 | -0.069, 0.269 | 0.245 | 0.223, 0.278 | 0.250 | 4263 | |
| Emotional stability personality trait | Beta | -0.067 | -0.263, 0.130 | 0.506 | 0.472, 0.534 | 0.503 | 4224 | |
| Extraversion personality trait | Beta | -0.042 | -0.243, 0.159 | 0.682 | 0.657, 0.716 | 0.687 | 4354 | |
| Frequency of doing exercise | Beta | -0.009 | -0.032, 0.014 | 0.450 | 0.443, 0.505 | 0.474 | 4270 | |
| Sleep duration (hours of sleep) | Beta | -0.016 | -0.047, 0.014 | 0.294 | 0.260, 0.317 | 0.288 | 3726 | |
| GCSE grades A-C | OR | 1.467 | 1.146, 1.877 | 0.005 | <0.001, 0.004 | <0.001 | 2360 | |
| GCSE grades D-G | OR | 1.007 | 0.914, 1.109 | 0.876 | 0.846, 0.889 | 0.869 | 2182 | |
| IQ | Beta | 0.138 | -0.293, 0.569 | 0.531 | 0.497, 0.559 | 0.528 | 3720 | |
| Sleep initiation | Beta | 0.015 | -0.019, 0.050 | 0.385 | 0.353, 0.414 | 0.383 | 3626 | |
| Sleep maintenance | Beta | -0.003 | -0.028, 0.022 | 0.804 | 0.813, 0.859 | 0.837 | 3418 | |
| Life events | Beta | -0.007 | -0.045, 0.031 | 0.733 | 0.690, 0.747 | 0.719 | 3376 | |
| Total number of outcomes tested = 46 |  |  |  |  |  |  |  | |

**Supplementary Table S8. Correlation between smoking, caffeine and alcohol GRS**

|  | **Smoking initiation GRS** | **Lifetime smoking GRS** | **Caffeine GRS** | **Alcohol GRS** |
| --- | --- | --- | --- | --- |
| **Smoking initiation GRS** | - | 0.35 | 0.01 | 0.08 |
|  | **Lifetime smoking GRS** | - | -0.01 | 0.02 |
|  |  | **Caffeine GRS** | - | 0.12 |

1. Tukey. Exploratory data analysis. Biometric Journal. 1981;5.
2. Farrow, A., Shea, K. M., & Little, R. E. (1998). Birthweight of term infants and maternal occupation in a prospective cohort of pregnant women. The ALSPAC Study Team. *Occupational and environmental medicine*, *55*(1), 18–23. doi:10.1136/oem.55.1.18
3. The Strengths and Difficulties Questionnaire
4. The Development and Well-Being Assessment
5. The Short Mood and Feelings Questionnaire
6. Colin, D., Jean, G., & Patrick, F. (2010). Traits contributing to the autistic spectrum. *Plos One,* *5*(9). doi:10.1371/journal.pone.0012633
7. The Wechsler-Intelligence Scale
8. The Revised Clinical Interview Schedule
9. The Mood and Feelings Questionnaire
10. Easey, K.E., Mars, B., Pearson, R. et al. Eur Child Adolesc Psychiatry (2019) 28: 1079. https://doi.org/10.1007/s00787-018-1266-1
11. The psychosis-like symptoms measure
12. The International Personality Item Pool
13. Wechsler abbreviated scale of Intelligence
14. Alcohol Use Disorders Identification Test
15. Edinburgh Postnatal Depression Scale
16. Interpersonal Sensitivity Measure
17. The Crown Crisp Experiential Index (anxiety sub-scale)
18. The Karolinska Scale of Personality
